# The genetic background of hydrocephalus in a population-based cohort: implication of ciliary involvement

**DOI:** 10.1101/2022.04.11.22273725

**Authors:** Tina N. Munch, Paula L. Hedley, Christian M. Hagen, Marie Bækvad-Hansen, Frank Geller, Jonas Bybjerg-Grauholm, Merete Nordentoft, Anders D. Børglum, Thomas M. Werge, Mads Melbye, David M. Hougaard, Lars A. Larsen, Søren T. Christensen, Michael Christiansen

## Abstract

**Background:** Hydrocephalus is one of the most common congenital disorders of the central nervous system and often displays psychiatric co-morbidities, in particular autism spectrum disorder. The disease mechanisms behind hydrocephalus are complex and not well understood, but some association with dysfunctional cilia in the brain ventricles and subarachnoid space has been indicated. A better understanding of the genetic aetiology of hydrocephalus, including the role of ciliopathies, may bring insights into a potentially shared genetic aetiology. In this population-based case-cohort study we, for the first time, investigated variants of postulated hydrocephalus candidate genes. Using this data, we aimed to investigate potential involvement of the ciliome in hydrocephalus and describe genotype-phenotype associations with autism spectrum disorder, identified in the iPSYCH cohort.

**Methods:** One-hundred and twenty-one hydrocephalus candidate genes were screened in a whole-exome sequenced sub cohort of the iPSYCH study, comprising 72 hydrocephalus patients and 4,181 background population controls. Candidate genes containing high-impact variants of interest were systematically evaluated for their involvement in ciliary function and autism spectrum disorder.

**Results:** The median age at diagnosis for the hydrocephalus patients was 0 years of age (range 0-27 years), the median age at analysis was 22 years (11-35 years), and 70.5 % were males. Median age for controls was 18 years (range 11-26 years) and 53.3 % were males. Fifty-two putative hydrocephalus-associated variants in 34 genes, were identified in 42 patients (58.3 %). In hydrocephalus cases, we found increased, but not significant, enrichment of high-impact protein altering variants (OR 1.51, 95% CI 0.92-2.51, p = 0.096), which was driven by a significant enrichment of rare protein truncating variants (OR 2.71, 95% CI 1.17-5.58, p = 0.011). Fourteen of the genes with high-impact variants are part of the ciliome, whereas another six genes affect cilia-dependent processes during neurogenesis. Furthermore, 15 of the 34 genes with high-impact variants and three of eight genes with protein truncating variants were associated with autism spectrum disorder.

**Conclusions:** Because symptoms of other diseases may be neglected or masked by the hydrocephalus-associated symptoms and identification of co-morbidities may be of clinical significance, we suggest that patients with congenital hydrocephalus undergo clinical genetic assessment with respect to ciliopathies and autism spectrum disorder. Our results point to the significance of hydrocephalus as a ciliary disease in some cases. Future studies in brain ciliopathies may not only reveal new insights into hydrocephalus, but also, brain disease in the broadest sense, given the essential role of cilia for neurodevelopment.

## Introduction

Hydrocephalus is a complex and heterogenic condition in which the volume of intracranial CSF is excessive relative to the brain volume due to different mechanisms encompassing 1) structural blockage of CSF circulation, 2) altered permeability of the ependymal lining of the ventricular system and choroid plexus, and 3) impaired neurodevelopment. At the same time, it is one of the most common congenital disorders of the central nervous system affecting 1.08 in 1,000 live-born children, when all subtypes are included.^1^ At the time of writing, 100+ candidate genes for hydrocephalus have been described in referred patients and animal studies,^2-5^ including eight genes known to exhibit a Mendelian pattern of inheritance, namely two X-linked genes; *L1CAM* and *AP1S2*, and six autosomal genes; *CCDC88C, MPDZ, TRIM71, SMARCC1, PTCH1*, and *SHH*.^2-5^ There are no findings from genome-wide association studies, which is probably due to difficulties to identify sufficient case numbers to identify moderate or small effect sizes, and the situation is further complicated by the heterogenous presentations of hydrocephalus. Several of the aforementioned genes are known to play important roles in early brain development, whereas others lead to impaired CSF dynamics, but overall, the understanding of the actual disease mechanisms leading to hydrocephalus at a molecular genetic level is still in a burgeoning stage.^3, 5-8^

Emerging evidence suggests a critical function of cilia in neurodevelopmental and psychiatric disorders.^9-12^ Cilia comprise a diverse group of antenna-like and microtubules-based organelles, which project from centrosomes or centrioles at the cell surface and play key roles in cell motility and/or signalling, in turn controlling development and function of many types of tissue and organs in our body.^11, 13^

Cilia in the brain have functional diversities and therefore ciliopathies may, by different mechanisms, lead to hydrocephalus. Motile cilia in the ependymal lining of the ventricular system conduct the wall-near CSF flow and together with the pulse generated bulk flow ensures circulation between the compartments of the ventricular system and further the flow through the outlets of the fourth ventricle to the subarachnoid space and further facilitating the circulation to the venous sites of CSF absorption.^14, 15^ Motile ciliopathies can lead to hydrocephalus due to impaired circulation and subsequent accumulation of CSF.^16, 17^ Furthermore, motile ciliopathies may impair the transport of signaling molecules and cells necessary for normal neurodevelopment.^15^ Another suggested mechanism is that decreased CSF flow impairs the patency of the different outlets of the ventricular system thereby causing obstructive hydrocephalus due to, for instance, stenosis of the Sylvian aqueduct that connect the third and fourth ventricle.^18, 19^

The immotile sensory/primary cilia in the ventricular lining interact with subventricular stem cells and play crucial roles in their activation, maturation, and migration due to the roles of primary cilia in Wnt- and sonic hedgehog signaling.^10, 20, 21^ The impaired formation of the brain parenchyma and/or structural abnormalities of the ventricular system can lead to hydrocephalus as well as typical associated abnormalities such as dys- or agenesia of the corpus callosum, decreased cortical development, posterior fossa abnormalities and associated cystic formations.^20, 21^ The choroid plexus cilia appear to only have transient motility during the perinatal period,^22, 23^ whereafter they are involved in regulating ion transport and CSF production.^24^ Impaired function may lead to hydrocephalus due to overproduction of CSF but without impairment of neurodevelopment.^24-26^

Ciliary genes encode structural proteins in the cilium, proteins directly involved in ciliary function, including the signalling pathways compartmentalised into the ciliary organelle, or genes involved in maintaining ciliary function or structural integrity within the cell. Understanding the genetic disease mechanisms leading to hydrocephalus may also provide an explanation of the strong association between hydrocephalus and autism spectrum disorder that has been found in another sub study of the Initiative for Integrative Psychiatric Research (iPSYCH) cohort,^27^ as well as in other populations.^28-30^ Access to the iPSYCH cohort data enabled us to conduct this population-based case-cohort study investigating virtually all postulated hydrocephalus candidate genes (121) in 72 whole-exome sequenced hydrocephalus cases for putatively hydrocephalus-causing variants, compared with 4,181 exome-sequenced background population controls without psychiatric disorders, hydrocephalus, or any of the neurologic conditions preselected by the iPSYCH group. Furthermore, we explored the involved disease mechanisms with specific focus on potential involvement of the ciliome as a mechanism of disease, and whether the involved genes previously have been associated with autism spectrum disorder.

## Materials and methods

### Data sources

This register-based cohort study is a sub study of iPSYCH^31^ and combines data from The Danish National Patient Register,^32^ The Danish Neonatal Screening Biobank,^33^ and the Danish Psychiatric Central Research Register,^34^ using the unique personal identification number from the Danish Civil Registration System.^35^

### Source population – the iPSYCH cohort

All members of the iPSYCH cohort were born between 1981 – 2005 and the cohort was followed until December 31, 2016. The cohort members fulfilled two basic requirements: 1) Being alive at one year of age, and 2) the mother was residing in Denmark holding a Danish Civil Registration number (CPR-number). The CPR number was used to link with the Danish National Patients Register as well as the Danish Psychiatric Central Register, in order to obtain information on diagnoses of the entire cohort. The Danish Civil Registration System covers the entire country, and it is continuously updated on vital status and migration. Reporting of diagnoses to the Danish National Patients and Psychiatric Central Register happens continuously and is mandatory for all public and private hospitals in Denmark. The cohort members were followed until one of the following events: migration, death, or end of follow-up December 31, 2016.

### Study population

Patients and controls were obtained from the iPSYCH cohort, which contains genetic and diagnostic information on 27,889 whole-exome sequenced Danes.^31^ Of these, 22,255 were cases who had received a psychiatric diagnosis, 429 had received a diagnosis with known psychiatric associations, and 5,205 background population controls were extracted from the randomly sampled iPSYCH population-based cohort.^36^ We identified 78 hydrocephalus cases and 4,259 background population controls without psychiatric disorders, hydrocephalus, or any of the neurological conditions preselected by the iPSYCH group (Huntington’s’ disease, Parkinson’s disease, epilepsy, migraine, birth defect in heart and major arteries or veins, syncope, and febrile seizures) registered during the study period from the exome-sequenced population in the iPSYCH cohort. No relatedness or stratification assessments were performed. Six hydrocephalus patients and 78 controls were removed from the analysis due to missingness (samples where > 5 % of variant sites were missing or failed). Out of the 72 remaining hydrocephalus cases, 50 were diagnosed with co-occurring autism spectrum disorder, 10 were diagnosed with other psychiatric disorders, six were diagnosed with epilepsy, and six individuals were from the background population. The hydrocephalus cases were compared to the remaining 4,181 background population controls. Figure 1 illustrates the selection steps of the study population.

**Figure 1.**
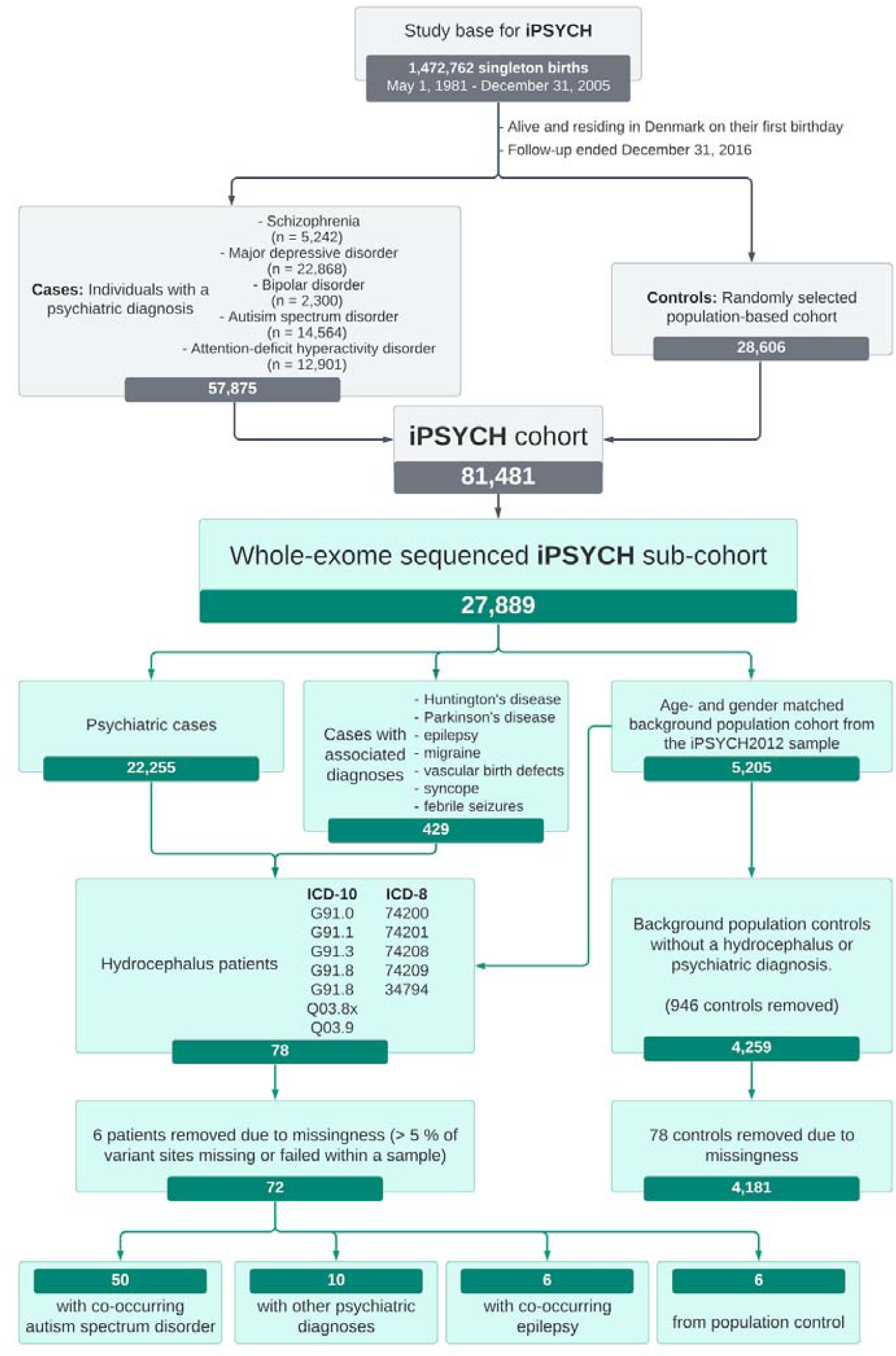
Flow chart of the study population. Grey boxes briefly indicate the distribution of cases and controls from the original iPSYCH case-cohort sample. The green boxes indicate the distribution of cases and controls in the present study utilizing the exome sequenced data from iPSYCH. The associated diagnoses represent neurologic conditions which have been associated with psychiatric disease within the iPSYCH cohort. The limit of missingness was >5 % of variant sites missing or failed within a sample. ASD: autism spectrum disorder.

### Ascertainment of hydrocephalus

The study base was linked, through the CPR number, to the National Patient Register. Hydrocephalus cases were identified using the following hydrocephalus diagnoses classified according to the two different versions of the International Classification of Diseases used during the study period. ICD-8: 74200, 74201, 74208, 74209, and 34794. ICD-10: G91.0, G91.1, G91.3, G91.8, G91.9, Q038x, and Q039.

### Selection and analysis of candidate genes

From the literature, we identified 121 genes where mutations had previously been implicated in hydrocephalus or were part of molecular pathways known to harbour hydrocephalus-causing genes^3, 5^ (Supplementary Table 1). Analyses of the 121 candidate genes were performed on the exomes from both cases and controls. DNA extraction and exome sequencing from dried blood spots have been described previously for this cohort.^31, 37^

### Quality control (QC) assessments

Filtering steps were adapted from Satterstrom et al (2021).^38^ Briefly, all variants were filtered as follows: 1) Variants with a GATK VQSR (variant quality score recalibration) label of “PASS” and a depth of coverage between 10 and 1000 were selected. 2) Low quality variants were removed; these variants were defined as: A) homozygous reference calls with a GQ < 25 or fewer than 90 % of reads supporting the reference allele call. B) Homozygous variant calls with a PL (HomRef) < 25 or fewer than 90 % of reads supporting the alternate allele call. C) Heterozygote calls with a PL (HomRef) < 25, fewer than 25 % reads supporting the alternate allele, fewer than 90 % informative reads, a probability of drawing the allele balance from a binomial distribution centred on 0.5 of less than 1e-9, or a location where the sample should be hemizygous (e.g., calls on the X chromosome outside the pseudo-autosomal region in a male). D) Y-chromosome calls (outside the pseudo-autosomal region) in a female. 3) Variants with a call rate below 80 % were removed.

As the sequence alignment map format was not accessible due to iPSYCH’s adherence to the American College of Medical Genetics recommendations for reporting incidental findings,^39^ indirect estimations of gene-specific exon sequencing coverage were performed using allele frequency (AF) from gnomAD v2.1.1 and the study population. Briefly, the gnomAD minor AF (MAF) for variants identified in samples of Non-Finnish European ancestry in the gnomAD controls subset (controls_AF_nfe) were compared to the MAF in variants identified in the study population. They were found to be highly correlated (r2 = >0.977, p = 0) for AF > 0.002. The number of expected identified variants per gene were defined as exon variants in gnomAD with controls_AF_nfe >= 0.002 (Corresponding to ∼13 alleles after adjusting for minimum accepted genotype call-rate (0.8)). The probability of not-having an observation at an included site were estimated to be P < 9.2E-7.

Samples with missing genotype calls Z-score > 3 or < -3 were excluded, six cases and 78 controls were removed, leaving 72 cases and 4,181 controls. Of the remaining, included samples the missingness per sample was not statistically significant between cases and controls (P = 0.44/0.17 t-test/Wilcoxon). The percentage of variants with statistically significant (P < 0.05) case/control missingness rate was 1.6 %.

Variants were annotated using SnpEff GRCh37.p13.RefSeq using only protein coding transcripts and CADD v.1.4. All conflicting variant annotations according to GRCh37.p13.RefSeq were excluded. A total of 5,854 variants remained after QC.

As an estimate of case/control comparability the proportions of synonymous alleles (allelic-load) in cases and controls were compared using two proportions z-test with and without adjusting for missing genotype rate. For the non-adjusted the proportions were 3.2 % for cases and 3.15 % for controls (P = 0.12) and the adjusted 3.2% and 3.2 % (P = 0.13), respectively.

### Prediction and classification of high-impact variants

To discriminate between high- and low-impact variants we used CADD v.1.4 PHRED scored variants in the candidate genes. Variants reported in ClinVar (clinvar_20210814)^40^ and gnomAD (v2.1.1) were used to find the best performing (f1-score) PHRED score cut-off for each of the sequence ontology classes (stop-gained, stop-lost, splice-site, non-synonymous, in-frame, frame-shift, and canonical-splice). Low-impact variants were defined as: variants reported in ClinVar in which the review status was reported as either: “reviewed_expert_panel”, “criteria_provided, multiple_submitters, no_conflicts”, or “criteria_provided, single_submitter” and the clinical significance was reported as either: “benign”, “likely_benign”, in addition to gnomAD (v2.1.1) variants with MAF in all samples or controls_AF_nfe >0.001 and not present in ClinVar. High-impact variants were defined as: variants reported in ClinVar in which the review status was reported as either: “reviewed_expert_panel”, “criteria_provided, multiple_submitters, no_conflicts”, or “criteria_provided, single_submitter” and the clinical significance was reported as “pathogenic”.

For sequence ontology classes: non-synonymous, splice-site, and in-frame the PHRED cut-offs were found using a classification and regression trees (CART) algorithm (rpart v.4.1-15). For sequence ontology classes: canonical-splice, frame-shift, stop-gained, and stop-lost, too few low-impact variants were identified, consequently all variants of these classes were classified as high-impact. The performance and methods used for the high-/ low-impact classification of each sequence ontology class is shown in Supplementary Table 3. Additionally, predicted high-impact variants were reclassified as low-impact if the MAF in controls_AF_nfe from gnomAD (v2.1.1) exceeded 0.001 and variants present in ClinVar were reclassified according to their clinical significance status. Comparing cases with controls, there was a statistically significant difference of being called missing or failed in 0.8 % of the high-impact variants.

### Sensitivity analysis

To assess the uncertainty of the chosen CADD PHRED and AF cut-offs several sensitivity analyses were performed for non-synonymous, splice-site, in-frame and protein truncating variants (PTVs) (defined as canonical-splice, frame-shift, stop-lost, stop-gained). AF ranging from the minimum observed in the study population frequency (0.0001) to 0.1 (non-synonymous variants) and 0.01, (splice-site variants, in-frame variants, and PTVs), were assessed in 0.0005 intervals (Supplementary Table 3). For each AF cut-off the high-impact variant allele load, adjusted for missing genotype rate, was calculated and compared for cases and controls using Fisher’s exact test (Supplementary Table 4). Since all in-frame and splice-site variants occurred in control samples, the CADD PHRED cut-offs were assessed in non-synonymous variants only. PHRED range from 11 to 31.8 in 0.2325 intervals. For each PHRED cut-off the high-impact variant allele load, adjusted for missing genotype rate, was calculated, and compared for cases and controls using Fisher’s exact test (Supplementary Table 5).

### Burden analyses of high-impact variants

The difference between the number of carriers of high-impact variants among cases and controls was tested using Fisher’s exact test. Subsequent analyses tested the variant categories PTV and non-PTV.

To assess the uncertainty of the chosen CADD PHRED and AF cut-offs, a sensitivity analysis was performed for high-impact variants, as described above. Fisher’s Exact Test was used to test if there statistically significant difference between cases and controls for each AF - PHRED combination (Supplementary Table 5). All statistical analyses were performed in R 3.6.1.

### Phenotypes and mechanisms of disease

The pathogenicity of the candidate genes with high-impact variants was investigated using The Human Gene Mutation Database (HGMD),^41^ Online Mendelian Inheritance in Man (OMIM),^42^ and CiliaCarta.^43^ Protein structure/function consequences and evolutionary conservation of all variants were assessed by mutation taster.^44^ Furthermore, we conducted a systematic literature search in order to detect hydrocephalus phenotypes not yet described in the gene databases, and to investigate the role of the gene in ciliary function. Thus, PubMed was searched for the name of each of the screen-positive genes and/or the protein in combination with “hydrocephalus”, “ventriculomegaly”, “cilia”, “ciliopathies”, “autism”, and “ASD”.

## Data availability

The data that support the findings of this study are available from the corresponding author, upon reasonable request.

## Results

Among the 27,889 whole exome-sequenced individuals, we identified 78 patients diagnosed with hydrocephalus. Following quality control assessment six hydrocephalus patients were excluded from further study. The hydrocephalus patients were compared to 4,181 background population controls (Figure 1). The median age at diagnosis for the hydrocephalus patients was 0 years of age (range 0-27 years), the median age at analysis was 22 years (11-35 years), and 70.5 % were males. Median age for controls was 18 years (range 11-26 years) and 53.3 % were males. Screening of the exomes for 121 previously reported hydrocephalus candidate genes resulted in the identification of 52 high-impact variants in 34 susceptibility genes, which are presented in Table 1. Of these, nine were carrying protein-truncating variants (PTV’s): *B3GALNT2, CELSR2, CENPF, DAG1, IFT172, MTO1, NF1, PTCH1*, and *MTO1*. All but *MTO1* are genes involved in ciliary function. Detailed descriptions of all hydrocephalus-associated genes with high-impact variants are described in the supplementary material with regards to gene function, phenotypes as well as associations to ciliary function and ASD.

**Table 1.**
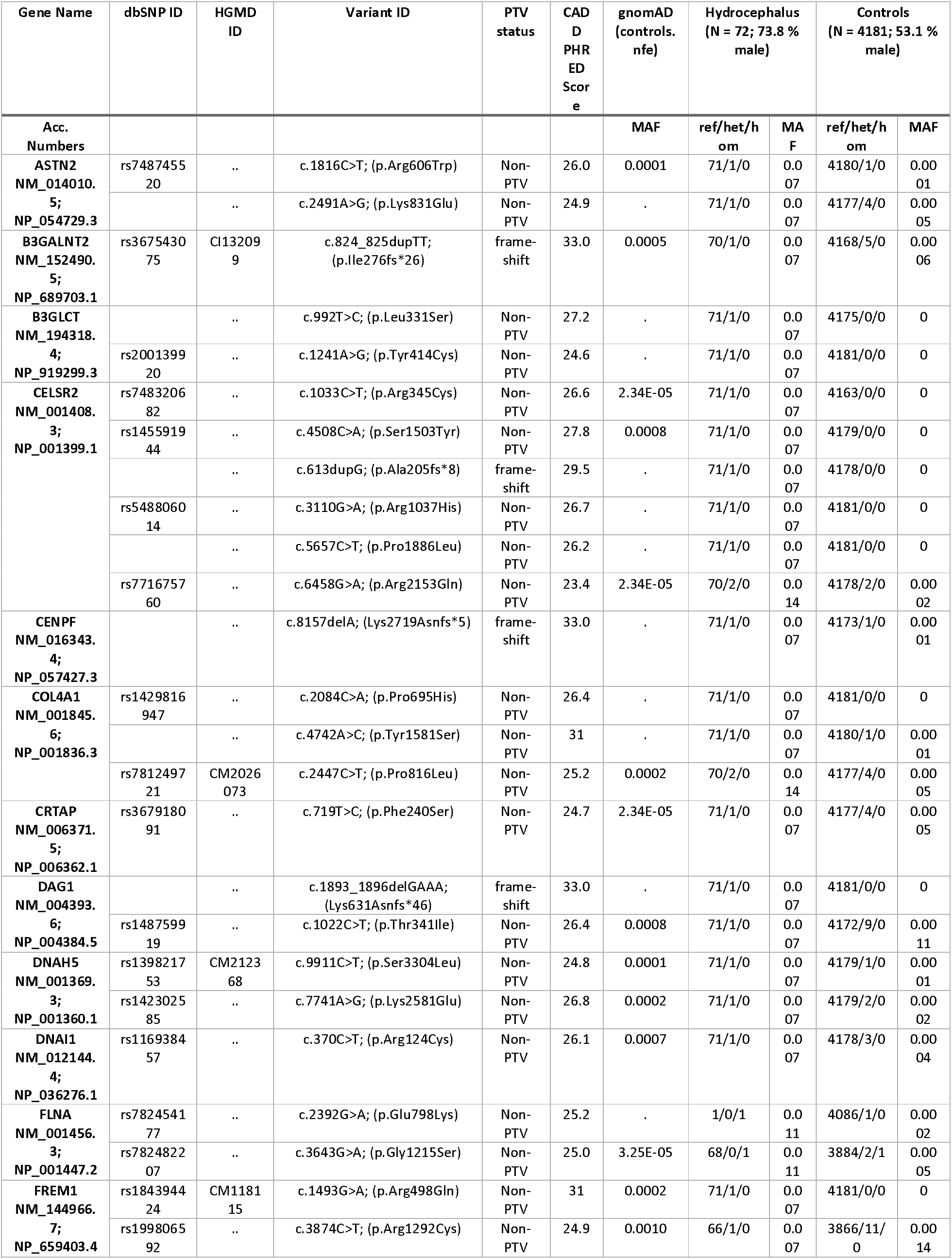
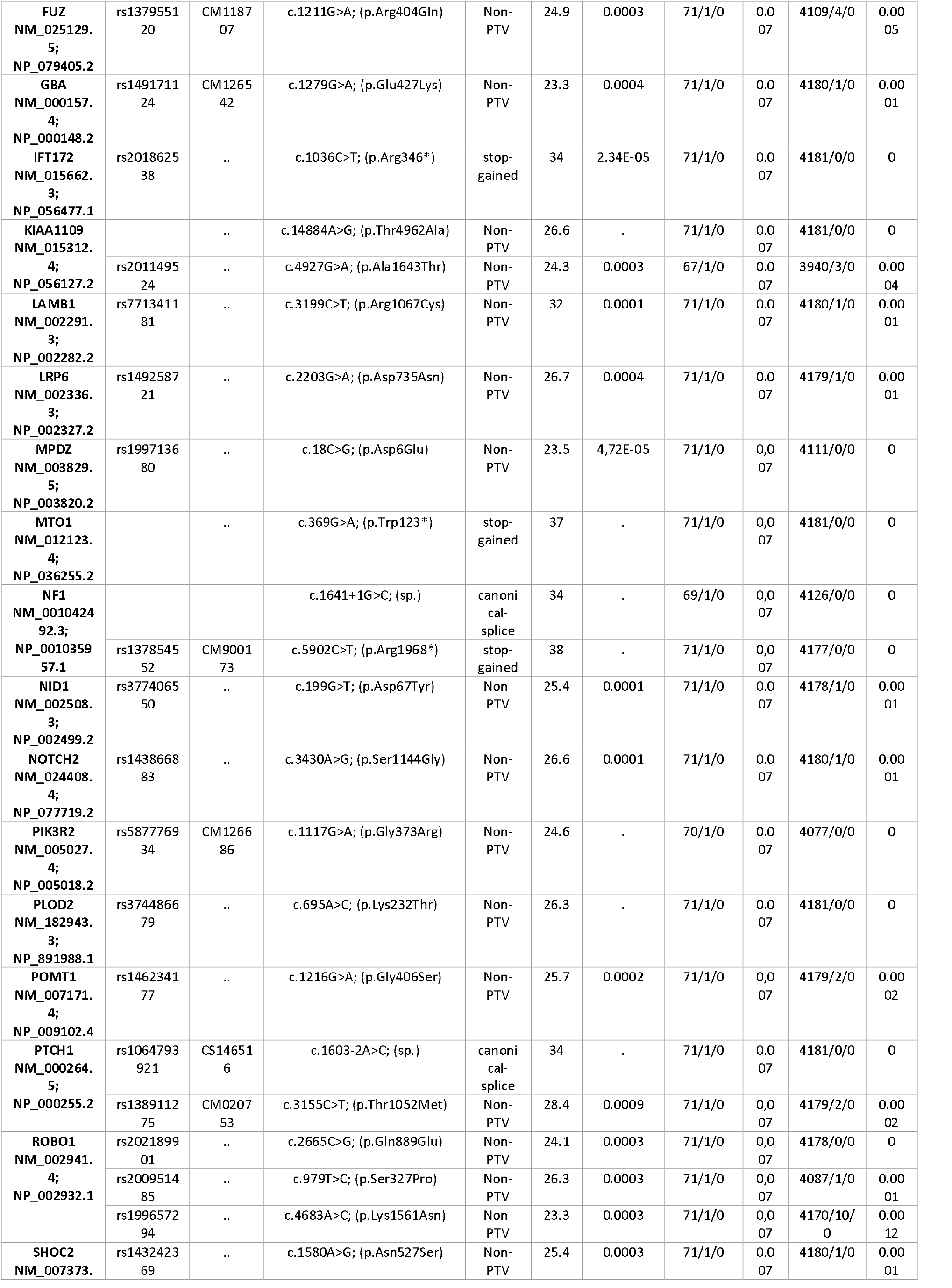
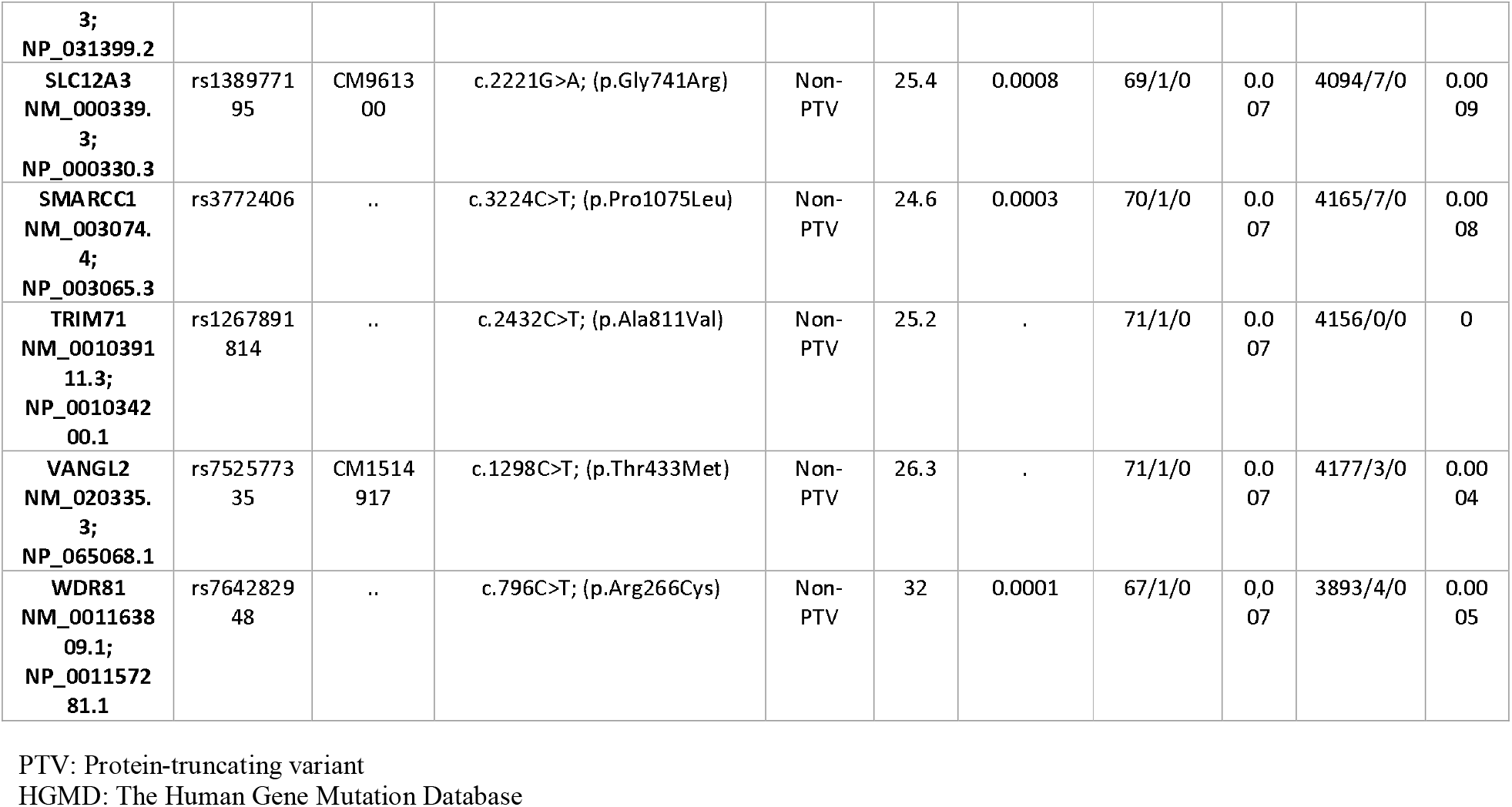
Overview of the 52 high-impact variants found in 34 of the 121 screened hydrocephalus-associated genes.

### Distribution of variants and burden analysis

Testing the carrier status for high-impact variants over all 121 genes of interest between hydrocephalus cases and controls showed an increased but not-significant number of high-impact variants among cases (OR 1.51, 95% CI 0.92-2.51, *P* = 0.096). Subsequent analysis of the variant sub-groups PTV’s and non-PTV’s, showed that the increase was mainly due to PTV’s (OR 2.71, 95% CI 1.17-5.58, *P* = 0.011, Table 2.

**Table 2.** Distribution of high-impact variants and protein truncating variants (PTVs) among the 121 genes screened in 72 hydrocephalus cases and 4, 181 background population controls. The results of burden tests are shown high-impact variants in total, as well as PTV- and non-PTV-high-impact variants.

| Variant type | Carriers by # Hits | Cases<br>N (%) | Controls<br>N (%) | P-value | OR | 95% CI |
| --- | --- | --- | --- | --- | --- | --- |
| **High-impact** | All carriers | 42 (58.3) | 2009 (48.1) | 0.096 | 1.51 | 0.92 - 2.51 |
| **High-impact** | 1 | 32 (44.4) | 1425 (34.1) | - | - | - |
| **High-impact** | 2 | 8 (11.1) | 456 (10.9) | - | - | - |
| **High-impact** | 3 | 2 (2.8) | 100 (2.4) | - | - | - |
| **High-impact** | 4 | 0 | 24 (0.6) | - | - | - |
| **High-impact** | 5 | 0 | 4 (0.1) | - | - | - |
| **PTVs** | All carriers | 9 (12.5) | 209 (5.0) | 0.011 | 2.71 | 1.17 - 5.58 |
| **PTVs** | 1 | 9 (12.5) | 200 (4.8) | - | - | - |
| **PTVs** | 2 | 0 | 9 (0.2) | - | - | - |
| **Non-PTVs** | All carriers | 33 (44) | 1800 (43.1) | 0.72 | 1.12 | 0.68 - 1.83 |

A detailed description of the distribution and types of variants found in the subset of 34 genes with high-impact variants in hydrocephalus cases is given in Supplementary Tables 6 and 7. No homozygote carriers, and two hemizygote carriers were identified among hydrocephalus cases, whereas one and four carriers, respectively, were identified among controls. Of the 52 high-impact variants carried by 42 of the hydrocephalus patients, nine were PTV’s, of which two were canonical splice site variants, four were frame-shift variants, and three were stop gained variants. All 43 non-PTVs were non-synonymous variants. As expected, given that the subset of genes is selected on high-impact variants in hydrocephalus cases, higher percentages were observed in the hydrocephalus cases.

### Genes with ciliary function

According to CiliaCarta (2019)^43^ and a literature search on each gene, we found that 14 of the 34 genes carrying high-impact variants had well-established ciliary function and another six genes were involved in cilia-dependent processes in neuronal formation. Among the eight genes carrying PTV’s, we found that four genes have well-described ciliary functions: *CELSR2* (primary and motile cilia),^45-47^ *CENPF* (ciliogenesis, microtubule dynamics),^43, 48^ *IFT172* (primary and motile cilia, intraflagellar transport),^43, 49^ *PTCH1* (primary and motile cilia).^2, 50^ Another three genes carrying PTV’s were found to be involved in processes regulated by primary cilia in the brain:*B3GALNT2, DAG1*, and *NF1*.

*B3GALNT2* encodes Beta-1,3-N-Acetylgalactosaminyltransferase 2, which is involved in the glycosylation of alpha-dystroglycan (α-DG). Reduced glycosylation impairs the interaction of α-DG with extracellular matrix elements,^51^ which may disrupt the signalling functions of primary cilia, and lead to brain malformations.^52^ *DAG1* which encodes dystrophin-associated glycoprotein 1, which forms part of the dystrophin-associated protein complex (DAPC) consisting of dystroglycans, sarcoglycans, dystrobrevins and syntrophins. DAPC components are expressed and regulated during the neuronal or astrocytic differentiation of neural stem/progenitor cells.^53^ *NF1* which encodes neurofibromin 1, a tumour suppressor which is expressed largely in neuronal cells, it is known to associate with microtubules and is involved in several signaling pathways.^54^ The neurofibromatosis changes may link to the function of primary cilia, as these are required for the formation of neural stem cells located in the subventricular zones, primarily of the lateral ventricles in adults and the third ventricle and optic pathway in children as well as for neurogenesis through Hedgehog signalling.^55-57^

The remaining 11 genes with well-defined ciliary function carrying high-impact variants (non-PTV’s) encompassed: *DNAH5* (motile cilia),^43, 58^ *DNAI1* (motile cilia),^59, 60^ *FLNA* (ciliogenesis),^43, 61^ *FUZ* (ciliogenesis, primary and motile cilia),^62-64^ *LRP6* (primary cilia),^65, 66^ *MPDZ* (ciliogenesis, motile cilia),^67^ *NOTCH2* (primary cilia),^68,69^ *PIK3R2* (primary cilia, ciliogenesis),^70^ *SMARCC1* (primary cilia),^71,72^ *TRIM71*(motile cilia),^73^ *VANGL2* (primary and motile cilia, ciliogenesis)^43, 46, 74^ In Table 3, all the high-impact variant carrying genes with ciliary involvement are described and the genes with PTV’s are underlined.

**Table 3.**
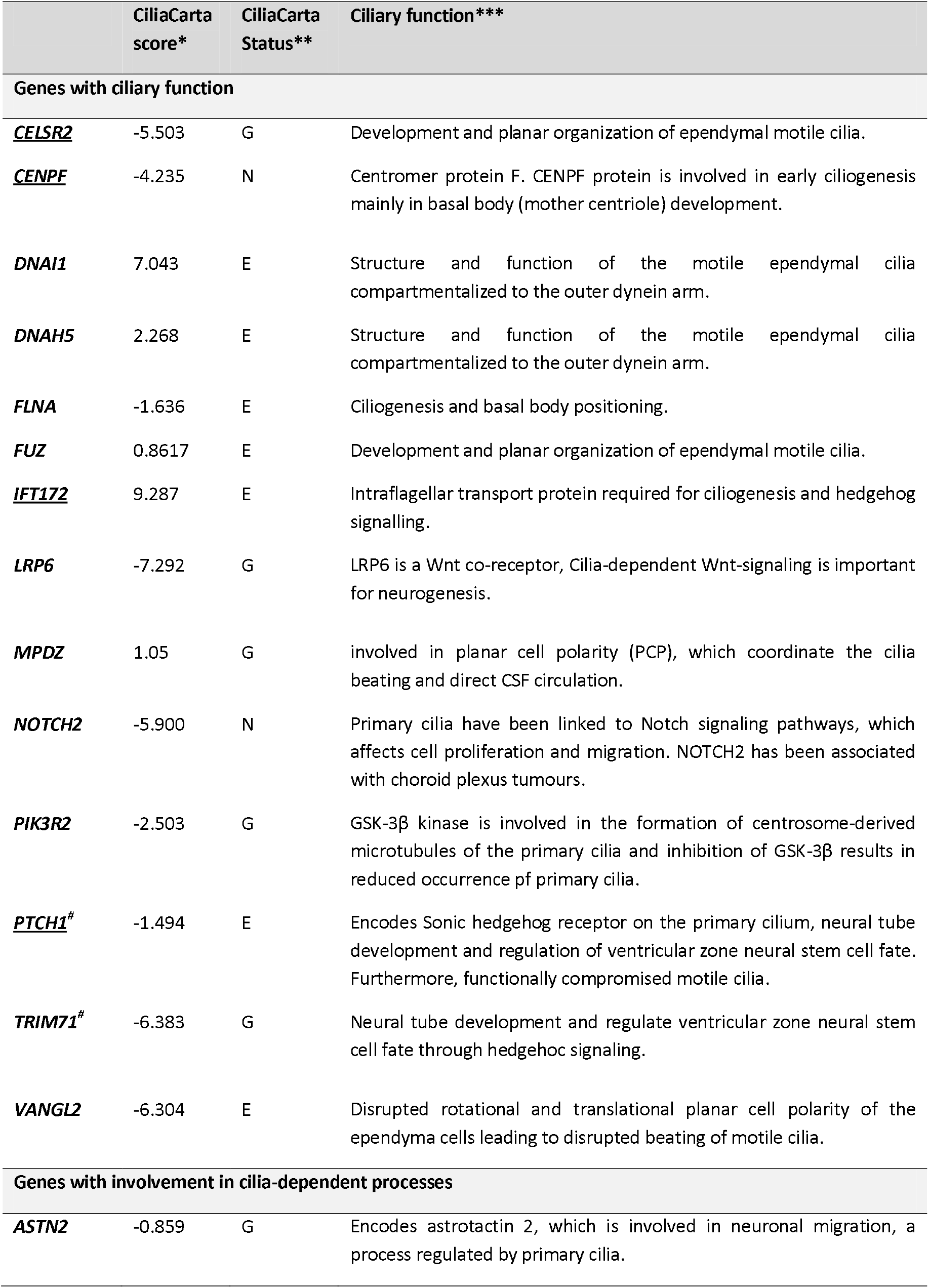
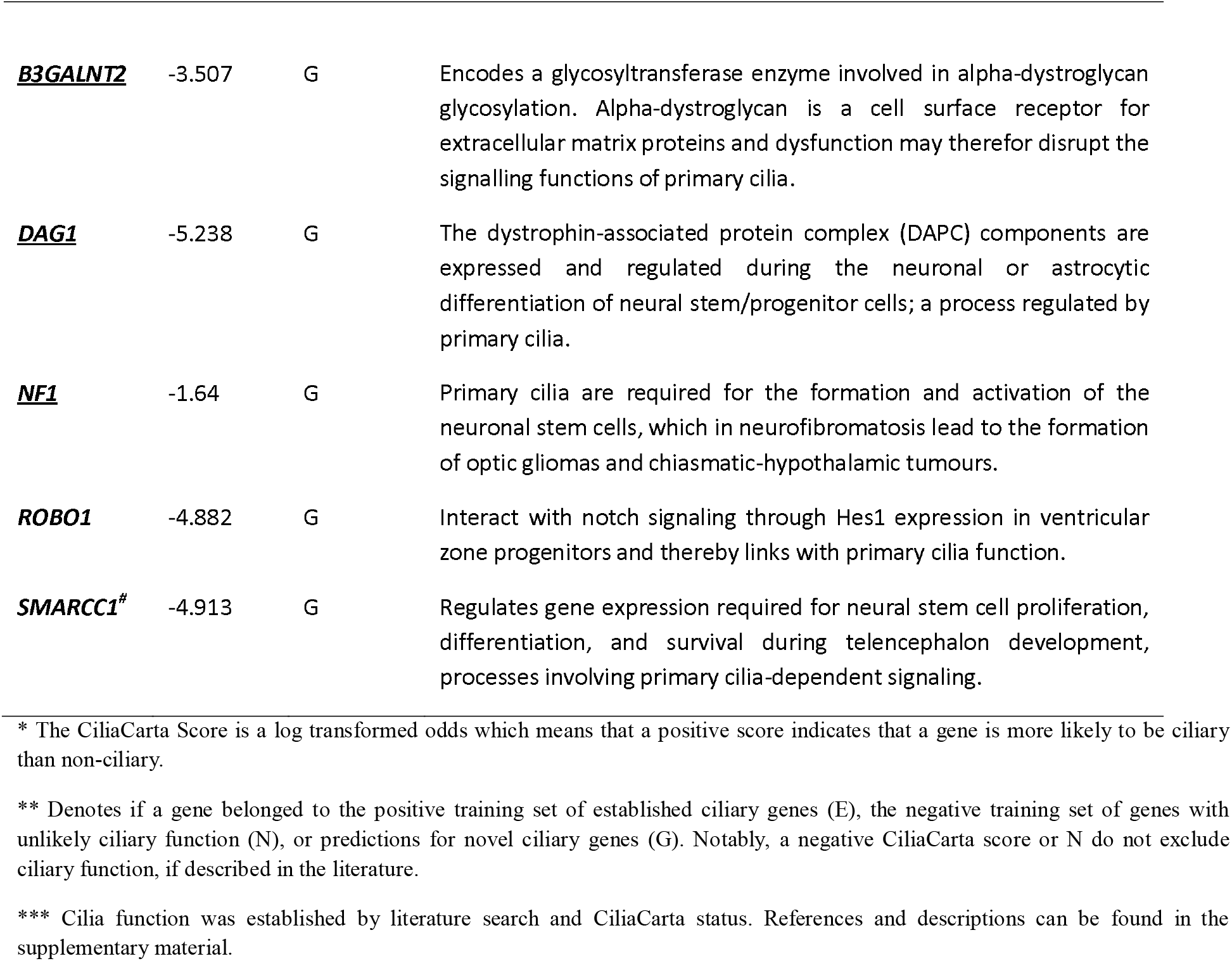
The subsets of 14 genes with ciliary function and the six genes with involvement in cilia-dependent processes, according to CiliaCarta and/or the literature among the 34 candidate genes associated with hydrocephalus. Gene carrying protein truncating variants have been underlined.

|  | CiliaCarta score* | CiliaCarta Status** | Ciliary function*** |
| --- | --- | --- | --- |
| **Genes with ciliary function** |  |  |  |
| <u>*CELSR2*</u> | -5.503 | G | Development and planar organization of ependymal motile cilia. |
| <u>*CENPF*</u> | -4.235 | N | Centromer protein F. CENPF protein is involved in early ciliogenesis mainly in basal body (mother centriole) development. |
| *DNAI1* | 7.043 | E | Structure and function of the motile ependymal cilia compartmentalized to the outer dynein arm. |
| *DNAH5* | 2.268 | E | Structure and function of the motile ependymal cilia compartmentalized to the outer dynein arm. |
| *FLNA* | -1.636 | E | Ciliogenesis and basal body positioning. |
| *FUZ* | 0.8617 | E | Development and planar organization of ependymal motile cilia. |
| <u>*IFT172*</u> | 9.287 | E | Intraflagellar transport protein required for ciliogenesis and hedgehog signalling. |
| *LRP6* | -7.292 | G | LRP6 is a Wnt co-receptor, Cilia-dependent Wnt-signaling is important for neurogenesis. |
| *MPDZ* | 1.05 | G | involved in planar cell polarity (PCP), which coordinate the cilia beating and direct CSF circulation. |
| *NOTCH2* | -5.900 | N | Primary cilia have been linked to Notch signaling pathways, which affects cell proliferation and migration. NOTCH2 has been associated with choroid plexus tumours. |
| *PIK3R2* | -2.503 | G | GSK-3β kinase is involved in the formation of centrosome-derived microtubules of the primary cilia and inhibition of GSK-3β results in reduced occurrence of primary cilia. |
| <u>*PTCH1*</u><sup>#</sup> | -1.494 | E | Encodes Sonic hedgehog receptor on the primary cilium, neural tube development and regulation of ventricular zone neural stem cell fate. Furthermore, functionally compromised motile cilia. |
| *TRIM71*<sup>#</sup> | -6.383 | G | Neural tube development and regulate ventricular zone neural stem cell fate through hedgehog signaling. |
| *VANGL2* | -6.304 | E | Disrupted rotational and translational planar cell polarity of the ependyma cells leading to disrupted beating of motile cilia. |
| **Genes with involvement in cilia-dependent processes** |  |  |  |
| *ASTN2* | -0.859 | G | Encodes astrotactin 2, which is involved in neuronal migration, a process regulated by primary cilia. |
| **<u>B3GALNT2</u>** | -3.507 | G | Encodes a glycosyltransferase enzyme involved in alpha-dystroglycan glycosylation. Alpha-dystroglycan is a cell surface receptor for extracellular matrix proteins and dysfunction may therefor disrupt the signalling functions of primary cilia. |
| **<u>DAG1</u>** | -5.238 | G | The dystrophin-associated protein complex (DAPC) components are expressed and regulated during the neuronal or astrocytic differentiation of neural stem/progenitor cells; a process regulated by primary cilia. |
| **<u>NF1</u>** | -1.64 | G | Primary cilia are required for the formation and activation of the neuronal stem cells, which in neurofibromatosis lead to the formation of optic gliomas and chiasmatic-hypothalamic tumours. |
| ***ROBO1*** | -4.882 | G | Interact with notch signaling through Hes1 expression in ventricular zone progenitors and thereby links with primary cilia function. |
| ***SMARCC1*<sup>#</sup>** | -4.913 | G | Regulates gene expression required for neural stem cell proliferation, differentiation, and survival during telencephalon development, processes involving primary cilia-dependent signaling. |

\* The CiliaCarta Score is a log transformed odds which means that a positive score indicates that a gene is more likely to be ciliary than non-ciliary.
\*\* Denotes if a gene belonged to the positive training set of established ciliary genes (E), the negative training set of genes with unlikely ciliary function (N), or predictions for novel ciliary genes (G). Notably, a negative CiliaCarta score or N do not exclude ciliary function, if described in the literature.
\*\*\* Cilia function was established by literature search and CiliaCarta status. References and descriptions can be found in the supplementary material.

### Genes associated with autism spectrum disorder

For 15 of the 34 genes with high-impact variants observed in the hydrocephalus cases (N = 20 carriers), there are reports in the literature for an involvement in ASD. Six patients carried PTV’s in three of these genes (*PTCH1, DAG1* and *NF1). PTCH1* is part of the ciliome, and *DAG1* and *NF1* are involved in cilia-dependent processes of neurogenesis. In total, 13 of the 15 high-impact variants (including all three PTV’s) were observed in patients with co-occurring ASD diagnosis. Proposed mechanisms of disease are summarized in Table 4 for the involved genes and elaborated in the supplementary material.

**Table 4.** Overview of the 15 hydrocephalus genes with high-impact variants carried by 20 patients, which were previously reported to be involved in ASD, according to phenotype descriptions in the literature. Genes carrying protein truncating variants have been underlined.

|  | Gene function | Suggested mechanism | Psychiatric comorbidity in carriers (N=20) |
| --- | --- | --- | --- |
| ***ASTN2*\*\*** | Encodes astrotactin 2, which is involved in neuronal migration, and is considered a ASD susceptibility gene. |  | ASD + affective disorder in two different carriers. |
| **<u>*DAG1*</u>\*\*** | Dystrophin-associated proteins complex are expressed and regulated during the neuronal or astrocytic differentiation of neural stem/progenitor cells | Impaired neuronal or astrocytic differentiation. | ASD + affective disorder in one carrier and schizophrenia in another. |
| ***FLNA*\*** | Filamin A has a crucial role in ciliogenesis and basal body positioning | Arrest of neuronal migration. | ASD in one carrier. |
| ***LAMB1*** | Encodes laminin, beta 1, an extracellular matrix protein. | Impaired axonal outgrowth and guidance. | ASD in one carrier. |
| ***MPDZ*\*** | Encodes multiple PDZ domain protein 1 (MUPP1), which is important for synaptic adhesion. Involved in planar cell polarity. | Impaired CASPR2-MUPP1-GPR37 complex on the dendrites is associated with one of the pathogeneses of ASD. | ASD + affective disorder in one carrier. |
| **<u>*NF1*</u>\*\*** | Formation of neural stem cells located in the subventricular zones, primarily of the lateral ventricles in adults and the third ventricle and optic pathway in children. | Impaired neurogenesis through hedgehog signalling. | ASD in one carrier and<br>ASD + ADHD in another. |
| ***NOTCH2*\*** | Encodes the notch receptor 2 and is thereby essential for notch signalling. | Impaired cortical neurogenesis. | ASD + ADHD in one carrier. |
| ***PIK3R2*\*** | Regulatory subunit of the PI3K-AKT-mTOR pathway, modulates GSK-3β activity. | Associated with brain overgrowth. GSK-3β kinase is involved in the formation of centrosome-derived microtubules of the primary cilia and inhibition of GSK-3β results in reduced occurrence of primary cilia. | ASD + affective disorder in one carrier. |
| ***PLOD2*** | Regulates the expression of downstream epithelial-mesenchymal transition-associated regulators in the PI3K/AKT signalling pathway. | ASD, together with skeletal abnormalities, has been associated with the *PLOD2* mutations. | ASD + affective disorder in one carrier. |
| ***POMT1*** | Encodes protein-O-mannosyltransferase 1, essential for alpha-dystroglycan (alpha-DG) glycosylation. | Impaired neuronal migration. | Affective disorder + schizophrenia in one carrier. |
| **<u>*PTCH1*</u>\*** | Encodes Patched 1, the Sonic hedgehog receptor | Impaired neural tube development and regulation of ventricular zone neural stem cell fate. | ASD + affective disorder in one carrier, ASD in another. |
| ***ROBO1*\*\*** | Regulates Hes1, a major repressor of Notch signalling. | *ROBO1* mutations interferes with early neural differentiation. | ASD in one carrier and<br>ADHD in another. |
| ***SHOC2*** | Encodes leucine-rich repeat scaffold protein, which is part of the RAS/MAPK pathway | Impaired neurogenesis. | ASD + ADHD in one carrier. |
| ***SMARCC1*\*\*** | Encodes an ATP-dependent chromatin remodeler that regulates gene expression required for neural stem cell proliferation. | Impaired neural stem cell proliferation, differentiation, and survival during telencephalon development. | Background population control. |
| ***VANGL2*\*** | Neural tube closure, PCP signalling, and beating of motile ependymal cilia. Involved in Wnt signalling. | Possibly affected interaction between Vangl2 and another PCP protein, the Prickle 2. | Background population control. |

\* Belongs to the ciliome (N=6)
\*\* Genes involved in cilia-dependent processes of neurogenesis (N=5)

## Discussion

In this population-based case-cohort study encompassing 72 hydrocephalus patients and 4,181 background population controls from the same birth cohort. Whole exome sequences from both cases and controls were screened for a panel of virtually all known candidate genes for hydrocephalus. Of the 121 investigated candidate genes, we identified 52 high-impact variants in 34 of the genes, carried by 42 of the hydrocephalus patients (58 %). The number of high-impact variants over all 121 genes of interest between hydrocephalus cases and controls was higher in hydrocephalus cases albeit not significant (OR 1.51, 95% CI 0.92-2.51, p = 0.096), whereas hydrocephalus cases were significantly enriched with the variant subgroup PTV’s (OR 2.71, 95% CI 1.17-5.58, p = 0.011). Interestingly, 20 of the 34 genes with high-impact variants (59 %), and 7 of the 8 (88 %) hydrocephalus genes with PTV’s, are either well-described part of the ciliome or are involved in cilia-dependent processes in neurogenesis.

### Hydrocephalus as a ciliopathy

The last two decades have seen a rapid increase in the number of genes involved in ciliogenesis and ciliary function, at the time of writing 302 genes with confirmed ciliary function according to CiliaCarta (2019),^43^ with many more reported in the literature. The functions of both primary and motile cilia are dependent upon interaction of several functional compartments in the organelle. Thus, elements in the basal body and transitional zone seem to be involved in pathways controlling more distal ciliary functioning. This also supports that modification of the function in a gene in one component may have significant effects on ciliary motor or sensory function. Ciliary function is very complex with function controlled by cytoplasmic signals as well as direct signals to specific locations on the cilium, e.g., sonic hedgehog-signaling through receptors on the “tip” of the primary cilium.^75, 76^ Furthermore, Wnt/β-catenin^65, 66^ and Notch signaling^77^ involves interaction with cilia.

Mutations in *CELSR2, PTCH1*, and *MPDZ* have been shown to impair ependymal flow, which in itself is believed to be one of the mechanisms of hydrocephalus formation.^19^ Furthermore, MRI-based computational fluid dynamic simulation in a human demonstrated that wall-near CSF flow is controlled by motility of ependymal cilia, whereas the flow in the centre of the third ventricle and Sylvian aqueduct are the result of macro-scale ventricular wall motion and choroid plexus pulsation.^14^ The direction of the CSF flow is determining for the direction of neuroblast migration, and thereby the organization of the brain tissue.^78^ Thus, deficient CSF flow may explain the co-existence of neurodevelopmental disorders and other brain pathologies in hydrocephalus. However, most of the screen-positive ciliary genes relate to the function of the primary cilium, which, by other mechanisms, also is crucial for neurodevelopment. Several of the screen-positive genes affect the three major signalling systems, Wnt/β-catenin, Sonic Hedgehog-, and Notch-signalling, which are associated with or compartmentalised to primary cilia.^13, 20, 79, 80^

*SMARCC1* and *TRIM 71* are two hydrocephalus associated genes with well-described phenotypes and involvement of primary and motile ciliary function, respectively. Whereas *SMARCC1* mutations cause aqueductal stenosis and with white matter volume reduction, as well as cardiac and skeletal abnormalities, patients with *TRIM71* mutations were prone to having communicating hydrocephalus with insufficient development of ependymal cells with motile cilia in the lining of the ventricular system.^2, 81^ A study in zebrafish revealed interaction between Smarcc1a and BBS6, which is involved in Bardet-Biedl syndrome. Bardet-Biedl syndrome serves as a study example on how several ciliary functions are affected; Bardet-Biedl syndrome mouse models involves ciliary intraflagellar transport, cilia maintenance, protein trafficking, and regulation of CSF production.^82, 83^ Corpus callosum abnormalities, complete septal agenesis or septal abnormalities, cerebellar tonsillar ectopia, developmental delay, epilepsy and malformations outside the CNS were additional common features in both *TRIM71* and *SMARCC1* mutation carriers.^81^ This illustrates the importance of both primary and motile cilia in neurogenesis. Rodríguez et al. suggest that the site of disrupted neurogenesis is of importance for the phenotype. The ventricular zone contains the multipotent radial glia/neural stem cells (NSCs) and the subventricular zone contains the rapidly proliferative neural precursor cells. According to Rodríguez et al., disruption of the ventricular zone of the Sylvian aqueduct leads to aqueductal stenosis, while disruption of the ventricular zone of telencephalon impairs neurogenesis.^84^

*CELSR2*, and *MPDZ* are involved in planar cell polarity (PCP), which together with intercellular junctional complexes, form the tissue structure and coordinated processes across epithelial sheets. In the ependymal cells with multiple cilia, rotational and translational PCP coordinate the cilia beating and direct CSF circulation. Thus, PCP is essential for ciliary function and disruption results in ciliopathies and hydrocephalus.^47, 85, 86^ Studies of Mpdz-deficient mice showed that expression of the interacting PCP protein Pals1 was diminished and barrier integrity got progressively lost leading to enhanced permeability of the choroid plexus.^26, 67, 87^ According to Feldner et al, the subsequent ependymal denudation was accompanied by reactive astrogliosis, which caused hydrocephalus due to aqueductal stenosis.^67^

The primary cilium is involved in integrating signals from the extracellular matrix and these interactions are significant for neuronal migration.^88^ Defective interaction with the extracellular matrix also interferes with Sonic Hedgehog and Notch signalling.^89, 90^ Therefore, several of the genes with PTVs may indirectly affect cilia-dependent processes in neurogenesis: *B3GALNT2* encodes an enzyme essential for the correct glycosylation of α-DG, which impairs the interaction of α-DG with extracellular matrix elements,^51, 91-95^ possibly disrupting the signalling functions of primary cilia. The genes *DAG1*, and *NF1* are involved in the regulation of neuronal or astrocytic differentiation of neural stem/progenitor cells, processes regulated by primary cilia.^53, 56, 57^

### Co-occurring autism spectrum disorder

In total, 15 genes with high-impact variants (including all three PTV’s) were previously associated with hydrocephalus-autism spectrum disorder phenotypes in the literature. For 13 of these genes, we found that the carriers had co-occurring autism spectrum disorder. Thus, one could ask if our genetic findings simply reflected the high occurrence of autism spectrum disorder among the hydrocephalus cases (69%). But according to a sub analysis (not shown), the burden of hydrocephalus-associated high-impact variants among patients with autism spectrum disorder was similar to controls. Hence, the presence of autism spectrum disorder does not influence the aetiology of hydrocephalus. As presented in Table 4, hydrocephalus and neurodevelopmental disorders may share mechanisms of disease through similar affected signalling pathways. Disruption of neuronal migration and differentiation is a key element in the etiology behind neurodevelopmental disorders^96, 97^, processes, which per above, to a large extend involves primary cilia.

### Limitations and strengths

A limitation of the present study is that detailed clinical information was unavailable. It would have been very informative to be able to describe somatic co-morbidities, as cilia-related diseases are characterised by a very broad range of syndromic and non-syndromic phenotypes. Some of these manifestations, such as recurring upper airway infections and otitis media, are likely treated outside the hospital system, and therefore these co-diagnoses will not be registered in the Danish National Patients Register. Unfortunately, cilia-associated diseases have no distinct ICD-codes, not even primary ciliary dyskinesia. Another limitation is that the study only includes individuals that were alive at one year of age, which may have led to an underestimation of hydrocephalus. Furthermore, the availability of trios would have improved interpretation of our findings with respect to causality. We observed an overrepresentation of males (70.5 %) among the cases compared to the background population controls (53.3 %), supposedly explained by the overrepresentation of males in hydrocephalus and autism spectrum disorder,^98, 99^ which constitutes a potential confounder. Yet, X-linked genetic causes were not driving our findings as only one out of 34 genes, *FLNA*, were localized to the X chromosome. Ideally, the study would have been replicated in an independent set of sequenced individuals with hydrocephalus from the general population and an independent group of hydrocephalus patients with autism spectrum disorder, and we advocate for such studies in the future. Notably, this is the first population-based study in which we investigated the presence of virtually all postulated hydrocephalus candidate genes, which is an important strength and improves the generalisability. Furthermore, all diagnoses were obtained from the Danish nationwide registries which boast complete national coverage.

### Clinical and research perspectives

The introduction of clinical genetics with respect to ciliopathies has the potential to become a game-changer in the personalized treatment of hydrocephalus. Advances in surgical treatment makes highly individualized treatment possible, but failure rates remain extremely high (cumulative 1-year risks of 44-50 %).^100, 101^ By causing disturbed micro circulation, and in some cases increased protein levels in the CSF, ciliopathies may very well be the prevailing causes of these persistently high failure rates. If the presence of a ciliopathy is accounted for in the choice of surgical treatment, the failure rate may decrease. Future studies of associations between genetic profile and surgical outcome could elucidate these hypotheses. For instance, a motile ciliopathy caused by *CELSR2, MPDZ*, and *PTCH1* may be better alleviated by shunt treatment rather than endoscopic treatment, since the impaired CSF flow would maintain a “functional aqueductal stenosis” after endoscopic treatment with third ventriculostomy. In the same line of thinking, mutations in *MPDZ* may help explain why some children are prone to shunt obstruction since mutations may lead to high permeability of the choroid plexus epithelial cell monolayer and thereby abnormally high CSF protein levels, which can cause shunt obstruction through accumulation of proteins in the ventricular catheter or shunt valve.^26^ Preoperative identification of *MPDZ* mutations may direct the treatment towards endoscopic surgical treatment, whenever possible and meaningful, or treatment with shunt systems less prone to obstruction.

Understanding the role of ciliopathies in hydrocephalus may furthermore pave the way for personalised medical treatment targeted at restoring ciliary function, as exemplified by the use of lentiviral gene therapy in Primary ciliary dyskinesia.^102^ Another example is the use of melanin-concentrating hormone that may increase the ciliary beating frequency.^103^ Moreover, downstream platelet-derived growth factor receptor activation by lithium in a Bardet-Biedl syndrome mouse model reduced the ventricular volume.^104^

## Conclusion

Based on several lines of evidence, our results point to the significance of hydrocephalus as a ciliary disease. We suggest clinical genetic assessment of congenital hydrocephalus patients with respect to ciliopathies and genetic predisposition for autism spectrum disorder as identification of co-morbidities may be of great clinical significance for the individual patient as symptoms of other diseases may be neglected or masked by the hydrocephalus-associated symptoms. Future research in brain ciliopathies may reveal new insights into brain disease in the broadest sense, given the essential role of cilia for neurodevelopment.

## Data Availability

All data produced in the present study are available upon reasonable request to the corresponding author.

## Abbreviations

α-DG: alpha-dystroglycan
AF: allele frequency
CART: classification and regression trees
DAPC: dystrophin-associated protein complex
HGMD: The Human Gene Mutation Database
iPSYCH: Lundbeck Foundation Initiative for Integrative Psychiatric Research
MAF: minor allele frequency
OMIM: Online Mendelian Inheritance in Man
PCP: planar cell polarity
PTV: protein truncating variant
VQSR: variant quality score recalibration

## Acknowledgements

We would like to acknowledge the iPSYCH group (www.iPSYCH.au.dk) and the Danish National Biobank who contributed to the data used in this study.

## Patient consent

The study is a sub-study of the iPSYCH study, that was approved by the Scientific Ethics Committees of the Central Denmark Region (www.komite.rm.dk) (J.nr.: 1-10-72-287-12) and the Danish Data Protection Agency (www.datatilsynet.dk) (J.nr.: 2012-41-0110). Passive consent was obtained, in accordance with Danish Law nr. 593 of June 14, 2011, para 10. The Danish Neonatal Screening Biobank approved use of the dried blood spot samples stored in the Danish Neonatal Screening Biobank. Detailed information on governance and ethics in the iPSYCH cohort is available at the iPSYCH website (www.ipsych.au.dk).

## References

1. Schmidt LB, Corn G, Wohlfahrt J, Melbye M, Munch TN. School performance in children with infantile hydrocephalus: a nationwide cohort study. Clin Epidemiol. 2018;10:1721–1731. doi:10.2147/clep.S178757

2. Furey CG, Choi J, Jin SC, et al. De Novo Mutation in Genes Regulating Neural Stem Cell Fate in Human Congenital Hydrocephalus. Neuron. Jul 25 2018;99(2):302-314.e4. doi:10.1016/j.neuron.2018.06.019

3. Kousi M, Katsanis N. The Genetic Basis of Hydrocephalus. Annu Rev Neurosci. Jul 8 2016;39:409–35. doi:10.1146/annurev-neuro-070815-014023

4. Shaheen R, Sebai MA, Patel N, et al. The genetic landscape of familial congenital hydrocephalus. Ann Neurol. Jun 2017;81(6):890–897. doi:10.1002/ana.24964

5. Jin SC, Dong W, Kundishora AJ, et al. Exome sequencing implicates genetic disruption of prenatal neuro-gliogenesis in sporadic congenital hydrocephalus. Nat Med. Oct 19 2020;doi:10.1038/s41591-020-1090-2

6. Weller S, Gärtner J. Genetic and clinical aspects of X-linked hydrocephalus (L1 disease): Mutations in the L1CAM gene. Hum Mutat. 2001;18(1):1–12. doi:10.1002/humu.1144

7. Furey CG, Choi J, Jin SC, et al. De Novo Mutation in Genes Regulating Neural Stem Cell Fate in Human Congenital Hydrocephalus. Neuron. Jul 25 2018;99(2):302–314 e4. doi:10.1016/j.neuron.2018.06.019

8. Furey CG, Zeng X, Dong W, et al. Human Genetics and Molecular Mechanisms of Congenital Hydrocephalus. World Neurosurg. Nov 2018;119:441–443. doi:10.1016/j.wneu.2018.09.018

9. Liu S, Trupiano MX, Simon J, Guo J, Anton ES. The essential role of primary cilia in cerebral cortical development and disorders. Curr Top Dev Biol. 2021;142:99–146. doi:10.1016/bs.ctdb.2020.11.003

10. Suciu SK, Caspary T. Cilia, neural development and disease. Semin Cell Dev Biol. Feb 2021;110:34–42. doi:10.1016/j.semcdb.2020.07.014

11. Wallmeier J, Nielsen KG, Kuehni CE, et al. Motile ciliopathies. Nat Rev Dis Primers. Sep 17 2020;6(1):77. doi:10.1038/s41572-020-0209-6

12. Wilsch-Bräuninger M, Huttner WB. Primary Cilia and Centrosomes in Neocortex Development. Front Neurosci. 2021;15:755867. doi:10.3389/fnins.2021.755867

13. Anvarian Z, Mykytyn K, Mukhopadhyay S, Pedersen LB, Christensen ST. Cellular signalling by primary cilia in development, organ function and disease. Nat Rev Nephrol. Apr 2019;15(4):199–219. doi:10.1038/s41581-019-0116-9

14. Siyahhan B, Knobloch V, de Zélicourt D, et al. Flow induced by ependymal cilia dominates near-wall cerebrospinal fluid dynamics in the lateral ventricles. J R Soc Interface. May 6 2014;11(94):20131189. doi:10.1098/rsif.2013.1189

15. Olstad EW, Ringers C, Hansen JN, et al. Ciliary Beating Compartmentalizes Cerebrospinal Fluid Flow in the Brain and Regulates Ventricular Development. Curr Biol. Jan 21 2019;29(2):229-241.e6. doi:10.1016/j.cub.2018.11.059

16. Kumar V, Umair Z, Kumar S, Goutam RS, Park S, Kim J. The regulatory roles of motile cilia in CSF circulation and hydrocephalus. Fluids Barriers CNS. Jul 7 2021;18(1):31. doi:10.1186/s12987-021-00265-0

17. Vogel P, Read RW, Hansen GM, et al. Congenital hydrocephalus in genetically engineered mice. Vet Pathol. Jan 2012;49(1):166–81. doi:10.1177/0300985811415708

18. Ibañez-Tallon I, Gorokhova S, Heintz N. Loss of function of axonemal dynein Mdnah5 causes primary ciliary dyskinesia and hydrocephalus. Hum Mol Genet. Mar 15 2002;11(6):715–21. doi:10.1093/hmg/11.6.715

19. Ibanez-Tallon I, Pagenstecher A, Fliegauf M, et al. Dysfunction of axonemal dynein heavy chain Mdnah5 inhibits ependymal flow and reveals a novel mechanism for hydrocephalus formation. Hum Mol Genet. Sep 15 2004;13(18):2133–41. doi:10.1093/hmg/ddh219

20. Youn YH, Han YG. Primary Cilia in Brain Development and Diseases. Am J Pathol. Jan 2018;188(1):11–22. doi:10.1016/j.ajpath.2017.08.031

21. Park SM, Jang HJ, Lee JH. Roles of Primary Cilia in the Developing Brain. Front Cell Neurosci. 2019;13:218. doi:10.3389/fncel.2019.00218

22. Nonami Y, Narita K, Nakamura H, Inoue T, Takeda S. Developmental changes in ciliary motility on choroid plexus epithelial cells during the perinatal period. Cytoskeleton (Hoboken). Dec 2013;70(12):797–803. doi:10.1002/cm.21132

23. Narita K, Kozuka-Hata H, Nonami Y, et al. Proteomic analysis of multiple primary cilia reveals a novel mode of ciliary development in mammals. Biol Open. Aug 15 2012;1(8):815–25. doi:10.1242/bio.20121081

24. Banizs B, Pike MM, Millican CL, et al. Dysfunctional cilia lead to altered ependyma and choroid plexus function, and result in the formation of hydrocephalus. Development. Dec 2005;132(23):5329–39. doi:10.1242/dev.02153

25. D’Gama PP, Qiu T, Cosacak MI, et al. Diversity and function of motile ciliated cell types within ependymal lineages of the zebrafish brain. Cell Rep. Oct 5 2021;37(1):109775. doi:10.1016/j.celrep.2021.109775

26. Yang J, Simonneau C, Kilker R, et al. Murine MPDZ-linked hydrocephalus is caused by hyperpermeability of the choroid plexus. EMBO Mol Med. Jan 2019;11(1)doi:10.15252/emmm.201809540

27. Munch TN, Hedley PL, Hagen CM, et al. Co-occurring hydrocephalus in autism spectrum disorder: a Danish population-based cohort study. J Neurodev Disord. Apr 28 2021;13(1):19. doi:10.1186/s11689-021-09367-0

28. Lindquist B, Carlsson G, Persson EK, Uvebrant P. Behavioural problems and autism in children with hydrocephalus : a population-based study. Eur Child Adolesc Psychiatry. Jun 2006;15(4):214–9. doi:10.1007/s00787-006-0525-8

29. Shen MD, Kim SH, McKinstry RC, et al. Increased Extra-axial Cerebrospinal Fluid in High-Risk Infants Who Later Develop Autism. Biol Psychiatry. Aug 1 2017;82(3):186–193. doi:10.1016/j.biopsych.2017.02.1095

30. Shen MD, Nordahl CW, Li DD, et al. Extra-axial cerebrospinal fluid in high-risk and normal-risk children with autism aged 2-4 years: a case-control study. Lancet Psychiatry. Nov 2018;5(11):895–904. doi:10.1016/S2215-0366(18)30294-3

31. Pedersen CB, Bybjerg-Grauholm J, Pedersen MG, et al. The iPSYCH2012 case-cohort sample: new directions for unravelling genetic and environmental architectures of severe mental disorders. Mol Psychiatry. Jan 2018;23(1):6–14. doi:10.1038/mp.2017.196

32. Andersen TF, Madsen M, Jorgensen J, Mellemkjoer L, Olsen JH. The Danish National Hospital Register. A valuable source of data for modern health sciences. Dan Med Bull. Jun 1999;46(3):263–8.

33. Grauholm J, Khoo SK, Nickolov RZ, et al. Gene expression profiling of archived dried blood spot samples from the Danish Neonatal Screening Biobank. Mol Genet Metab. Nov 2015;116(3):119–24. doi:10.1016/j.ymgme.2015.06.011

34. Mors O, Perto GP, Mortensen PB. The Danish Psychiatric Central Research Register. Scand J Public Health. Jul 2011;39(7 Suppl):54–7. doi:10.1177/1403494810395825

35. Schmidt M, Pedersen L, Sørensen HT. The Danish Civil Registration System as a tool in epidemiology. Eur J Epidemiol. Aug 2014;29(8):541–9. doi:10.1007/s10654-014-9930-3

36. Pedersen CB, Bybjerg-Grauholm J, Pedersen MG, et al. The iPSYCH2012 case-cohort sample: new directions for unravelling genetic and environmental architectures of severe mental disorders. Mol Psychiatry. Jan 2018;23(1):6–14. doi:10.1038/mp.2017.196

37. Poulsen JB, Lescai F, Grove J, et al. High-Quality Exome Sequencing of Whole-Genome Amplified Neonatal Dried Blood Spot DNA. PLoS One. 2016;11(4):e0153253. doi:10.1371/journal.pone.0153253

38. Satterstrom FK, Walters RK, Singh T, et al. ASD and ADHD have a similar burden of rare protein-truncating variants. bioRxiv. 2018:277707. doi:10.1101/277707

39. Miller DT, Lee K, Chung WK, et al. ACMG SF v3.0 list for reporting of secondary findings in clinical exome and genome sequencing: a policy statement of the American College of Medical Genetics and Genomics (ACMG). Genet Med. Aug 2021;23(8):1381–1390. doi:10.1038/s41436-021-01172-3

40. https://ftp.ncbi.nlm.nih.gov/pub/clinvar/vcf_GRCh37/archive_2.0/2021/.

41. Stenson PD, Mort M, Ball EV, Shaw K, Phillips A, Cooper DN. The Human Gene Mutation Database: building a comprehensive mutation repository for clinical and molecular genetics, diagnostic testing and personalized genomic medicine. Hum Genet. Jan 2014;133(1):1–9. doi:10.1007/s00439-013-1358-4

42. Amberger JS, Bocchini CA, Schiettecatte F, Scott AF, Hamosh A. OMIM.org: Online Mendelian Inheritance in Man (OMIM®), an online catalog of human genes and genetic disorders. Nucleic Acids Res. Jan 2015;43(Database issue):D789–98. doi:10.1093/nar/gku1205

43. van Dam TJP, Kennedy J, van der Lee R, et al. CiliaCarta: An integrated and validated compendium of ciliary genes. PLoS One. 2019;14(5):e0216705. doi:10.1371/journal.pone.0216705

44. Schwarz JM, Cooper DN, Schuelke M, Seelow D. MutationTaster2: mutation prediction for the deep-sequencing age. Nat Methods. Apr 2014;11(4):361–2. doi:10.1038/nmeth.2890

45. Shaheen R, Szymanska K, Basu B, et al. Characterizing the morbid genome of ciliopathies. Genome Biol. Nov 28 2016;17(1):242. doi:10.1186/s13059-016-1099-5

46. Yamasaki M, Kanemura Y. Molecular Biology of Pediatric Hydrocephalus and Hydrocephalus-related Diseases. Neurol Med Chir (Tokyo). 2015;55(8):640–6. doi:10.2176/nmc.ra.2015-0075

47. Tissir F, Qu Y, Montcouquiol M, et al. Lack of cadherins Celsr2 and Celsr3 impairs ependymal ciliogenesis, leading to fatal hydrocephalus. Nat Neurosci. Jun 2010;13(6):700–7. doi:10.1038/nn.2555

48. Pfaltzgraff ER, Roth GM, Miller PM, Gintzig AG, Ohi R, Bader DM. Loss of CENP-F results in distinct microtubule-related defects without chromosomal abnormalities. Mol Biol Cell. Jul 1 2016;27(13):1990–9. doi:10.1091/mbc.E15-12-0848

49. Friedland-Little JM, Hoffmann AD, Ocbina PJ, et al. A novel murine allele of Intraflagellar Transport Protein 172 causes a syndrome including VACTERL-like features with hydrocephalus. Hum Mol Genet. Oct 1 2011;20(19):3725–37. doi:10.1093/hmg/ddr241

50. Eggenschwiler JT, Anderson KV. Cilia and developmental signaling. Annu Rev Cell Dev Biol. 2007;23:345–73. doi:10.1146/annurev.cellbio.23.090506.123249

51. Stevens E, Carss KJ, Cirak S, et al. Mutations in B3GALNT2 cause congenital muscular dystrophy and hypoglycosylation of alpha-dystroglycan. Am J Hum Genet. Mar 7 2013;92(3):354–65. doi:10.1016/j.ajhg.2013.01.016

52. Nickolls AR, Lee MM, Zukosky K, Mallon BS, Bönnemann CG. Human embryoid bodies as a 3D tissue model of the extracellular matrix and α-dystroglycanopathies. Dis Model Mech. Jun 26 2020;13(6)doi:10.1242/dmm.042986

53. Romo-Yáñez J, Rodríguez-Martínez G, Aragón J, et al. Characterization of the expression of dystrophins and dystrophin-associated proteins during embryonic neural stem/progenitor cell differentiation. Neurosci Lett. Sep 25 2020;736:135247. doi:10.1016/j.neulet.2020.135247

54. Trovó-Marqui AB, Tajara EH. Neurofibromin: a general outlook. Clin Genet. Jul 2006;70(1):1–13. doi:10.1111/j.1399-0004.2006.00639.x

55. Dahiya S, Lee DY, Gutmann DH. Comparative characterization of the human and mouse third ventricle germinal zones. J Neuropathol Exp Neurol. Jul 2011;70(7):622–33. doi:10.1097/NEN.0b013e31822200aa

56. Han YG, Spassky N, Romaguera-Ros M, et al. Hedgehog signaling and primary cilia are required for the formation of adult neural stem cells. Nat Neurosci. Mar 2008;11(3):277–84. doi:10.1038/nn2059

57. Tong CK, Han YG, Shah JK, Obernier K, Guinto CD, Alvarez-Buylla A. Primary cilia are required in a unique subpopulation of neural progenitors. Proc Natl Acad Sci U S A. Aug 26 2014;111(34):12438–43. doi:10.1073/pnas.1321425111

58. Roberts AJ, Kon T, Knight PJ, Sutoh K, Burgess SA. Functions and mechanics of dynein motor proteins. Nat Rev Mol Cell Biol. Nov 2013;14(11):713–26. doi:10.1038/nrm3667

59. Ostrowski LE, Yin W, Rogers TD, et al. Conditional deletion of dnaic1 in a murine model of primary ciliary dyskinesia causes chronic rhinosinusitis. Am J Respir Cell Mol Biol. Jul 2010;43(1):55–63. doi:10.1165/rcmb.2009-0118OC

60. Cho KJ, Noh SH, Han SM, et al. ZMYND10 stabilizes intermediate chain proteins in the cytoplasmic pre-assembly of dynein arms. PLoS Genet. Mar 2018;14(3):e1007316. doi:10.1371/journal.pgen.1007316

61. Adams M, Simms RJ, Abdelhamed Z, et al. A meckelin-filamin A interaction mediates ciliogenesis. Hum Mol Genet. Mar 15 2012;21(6):1272–86. doi:10.1093/hmg/ddr557

62. Gray RS, Abitua PB, Wlodarczyk BJ, et al. The planar cell polarity effector Fuz is essential for targeted membrane trafficking, ciliogenesis and mouse embryonic development. Nat Cell Biol. Oct 2009;11(10):1225–32. doi:10.1038/ncb1966

63. Zhang W, Taylor SP, Ennis HA, et al. Expanding the genetic architecture and phenotypic spectrum in the skeletal ciliopathies. Hum Mutat. Jan 2018;39(1):152–166. doi:10.1002/humu.23362

64. Takagishi M, Sawada M, Ohata S, et al. Daple Coordinates Planar Polarized Microtubule Dynamics in Ependymal Cells and Contributes to Hydrocephalus. Cell Rep. Jul 25 2017;20(4):960–972. doi:10.1016/j.celrep.2017.06.089

65. Bryja V, Červenka I, Čajánek L. The connections of Wnt pathway components with cell cycle and centrosome: side effects or a hidden logic? Crit Rev Biochem Mol Biol. Dec 2017;52(6):614–637. doi:10.1080/10409238.2017.1350135

66. Gupta GD, Coyaud É, Gonçalves J, et al. A Dynamic Protein Interaction Landscape of the Human Centrosome-Cilium Interface. Cell. Dec 3 2015;163(6):1484–99. doi:10.1016/j.cell.2015.10.065

67. Feldner A, Adam MG, Tetzlaff F, et al. Loss of Mpdz impairs ependymal cell integrity leading to perinatal-onset hydrocephalus in mice. EMBO Mol Med. Jul 2017;9(7):890–905. doi:10.15252/emmm.201606430

68. Fiddes IT, Lodewijk GA, Mooring M, et al. Human-Specific NOTCH2NL Genes Affect Notch Signaling and Cortical Neurogenesis. Cell. May 31 2018;173(6):1356-1369.e22. doi:10.1016/j.cell.2018.03.051

69. Grisanti L, Revenkova E, Gordon RE, Iomini C. Primary cilia maintain corneal epithelial homeostasis by regulation of the Notch signaling pathway. Development. Jun 15 2016;143(12):2160–71. doi:10.1242/dev.132704

70. Fokin Artem I, Zhapparova Olga N, Burakov Anton V, Nadezhdina Elena S. Centrosome-derived microtubule radial array, PCM-1 protein, and primary cilia formation. Protoplasma. Sep 2019;256(5):1361–1373. doi:10.1007/s00709-019-01385-z

71. Narayanan R, Pirouz M, Kerimoglu C, et al. Loss of BAF (mSWI/SNF) Complexes Causes Global Transcriptional and Chromatin State Changes in Forebrain Development. Cell Rep. Dec 1 2015;13(9):1842–54. doi:10.1016/j.celrep.2015.10.046

72. Kerr EN, Bhan A, Héon E. Exploration of the cognitive, adaptive and behavioral functioning of patients affected with Bardet-Biedl syndrome. Clin Genet. Apr 2016;89(4):426–433. doi:10.1111/cge.12614

73. Cuevas E, Rybak-Wolf A, Rohde AM, Nguyen DT, Wulczyn FG. Lin41/Trim71 is essential for mouse development and specifically expressed in postnatal ependymal cells of the brain. Front Cell Dev Biol. 2015;3:20. doi:10.3389/fcell.2015.00020

74. Sheng X, Gao S, Sheng Y, Xie X, Wang J, He Y. Vangl2 participates in the primary ciliary assembly under low fluid shear stress in hUVECs. Cell Tissue Res. Jan 2022;387(1):95–109. doi:10.1007/s00441-021-03546-0

75. Pala R, Alomari N, Nauli SM. Primary Cilium-Dependent Signaling Mechanisms. Int J Mol Sci. Oct 28 2017;18(11)doi:10.3390/ijms18112272

76. Stoufflet J, Chaulet M, Doulazmi M, et al. Primary cilium-dependent cAMP/PKA signaling at the centrosome regulates neuronal migration. Sci Adv. Sep 2020;6(36)doi:10.1126/sciadv.aba3992

77. Jiang Z, Zhou J, Qin X, et al. MT1-MMP deficiency leads to defective ependymal cell maturation, impaired ciliogenesis, and hydrocephalus. JCI Insight. May 7 2020;5(9)doi:10.1172/jci.insight.132782

78. Sawamoto K, Wichterle H, Gonzalez-Perez O, et al. New neurons follow the flow of cerebrospinal fluid in the adult brain. Science. Feb 3 2006;311(5761):629–32. doi:10.1126/science.1119133

79. Seeger-Nukpezah T, Golemis EA. The extracellular matrix and ciliary signaling. Curr Opin Cell Biol. Oct 2012;24(5):652–61. doi:10.1016/j.ceb.2012.06.002

80. Murdoch JN, Copp AJ. The relationship between sonic Hedgehog signaling, cilia, and neural tube defects. Birth Defects Res A Clin Mol Teratol. Aug 2010;88(8):633–52. doi:10.1002/bdra.20686

81. Jin SC, Dong W, Kundishora AJ, et al. Exome sequencing implicates genetic disruption of prenatal neuro-gliogenesis in sporadic congenital hydrocephalus. Nat Med. Nov 2020;26(11):1754–1765. doi:10.1038/s41591-020-1090-2

82. Leigh MW, Pittman JE, Carson JL, et al. Clinical and genetic aspects of primary ciliary dyskinesia/Kartagener syndrome. Genet Med. Jul 2009;11(7):473–87. doi:10.1097/GIM.0b013e3181a53562

83. Swiderski RE, Agassandian K, Ross JL, Bugge K, Cassell MD, Yeaman C. Structural defects in cilia of the choroid plexus, subfornical organ and ventricular ependyma are associated with ventriculomegaly. Fluids Barriers CNS. Oct 9 2012;9(1):22. doi:10.1186/2045-8118-9-22

84. Rodríguez EM, Guerra MM, Vío K, et al. A cell junction pathology of neural stem cells leads to abnormal neurogenesis and hydrocephalus. Biol Res. 2012;45(3):231–42. doi:10.4067/s0716-97602012000300005

85. Fuertes-Alvarez S, Maeso-Alonso L, Villoch-Fernandez J, et al. p73 regulates ependymal planar cell polarity by modulating actin and microtubule cytoskeleton. Cell Death Dis. Dec 5 2018;9(12):1183. doi:10.1038/s41419-018-1205-6

86. Kim SK, Shindo A, Park TJ, et al. Planar cell polarity acts through septins to control collective cell movement and ciliogenesis. Science. Sep 10 2010;329(5997):1337–40. doi:10.1126/science.1191184

87. Al-Jezawi NK, Al-Shamsi AM, Suleiman J, et al. Compound heterozygous variants in the multiple PDZ domain protein (MPDZ) cause a case of mild non-progressive communicating hydrocephalus. BMC Med Genet. Mar 2 2018;19(1):34. doi:10.1186/s12881-018-0540-x

88. Banda E, McKinsey A, Germain N, Carter J, Anderson NC, Grabel L. Cell polarity and neurogenesis in embryonic stem cell-derived neural rosettes. Stem Cells Dev. Apr 15 2015;24(8):1022–33. doi:10.1089/scd.2014.0415

89. Dave RK, Ellis T, Toumpas MC, et al. Sonic hedgehog and notch signaling can cooperate to regulate neurogenic divisions of neocortical progenitors. PLoS One. Feb 17 2011;6(2):e14680. doi:10.1371/journal.pone.0014680

90. Lehtinen MK, Walsh CA. Neurogenesis at the brain-cerebrospinal fluid interface. Annu Rev Cell Dev Biol. 2011;27:653–79. doi:10.1146/annurev-cellbio-092910-154026

91. Uribe ML, Haro C, Ventero MP, Campello L, Cruces J, Martín-Nieto J. Expression pattern in retinal photoreceptors of POMGnT1, a protein involved in muscle-eye-brain disease. Mol Vis. 2016;22:658–73.

92. Stevens E, Carss KJ, Cirak S, et al. Mutations in B3GALNT2 cause congenital muscular dystrophy and hypoglycosylation of α-dystroglycan. Am J Hum Genet. Mar 7 2013;92(3):354–65. doi:10.1016/j.ajhg.2013.01.016

93. Hehr U, Uyanik G, Gross C, et al. Novel POMGnT1 mutations define broader phenotypic spectrum of muscle-eye-brain disease. Neurogenetics. Nov 2007;8(4):279–88. doi:10.1007/s10048-007-0096-y

94. Pozzi A, Yurchenco PD, Iozzo RV. The nature and biology of basement membranes. Matrix Biol. Jan 2017;57-58:1–11. doi:10.1016/j.matbio.2016.12.009

95. Uribe ML, Martín-Nieto J, Quereda C, et al. Retinal Proteomics of a Mouse Model of Dystroglycanopathies Reveals Molecular Alterations in Photoreceptors. J Proteome Res. Jun 4 2021;20(6):3268–3277. doi:10.1021/acs.jproteome.1c00126

96. Valiente M, Marín O. Neuronal migration mechanisms in development and disease. Curr Opin Neurobiol. Feb 2010;20(1):68–78. doi:10.1016/j.conb.2009.12.003

97. Lionel AC, Tammimies K, Vaags AK, et al. Disruption of the ASTN2/TRIM32 locus at 9q33.1 is a risk factor in males for autism spectrum disorders, ADHD and other neurodevelopmental phenotypes. Hum Mol Genet. May 15 2014;23(10):2752–68. doi:10.1093/hmg/ddt669

98. Munch TN, Rostgaard K, Rasmussen ML, Wohlfahrt J, Juhler M, Melbye M. Familial aggregation of congenital hydrocephalus in a nationwide cohort. Brain. Aug 2012;135(Pt 8):2409–15. doi:10.1093/brain/aws158

99. Schendel DE, Thorsteinsson E. Cumulative Incidence of Autism Into Adulthood for Birth Cohorts in Denmark, 1980-2012. Jama. Nov 6 2018;320(17):1811–1813. doi:10.1001/jama.2018.11328

100. Zaben M, Manivannan S, Sharouf F, et al. The efficacy of endoscopic third ventriculostomy in children 1 year of age or younger: A systematic review and meta-analysis. Eur J Paediatr Neurol. Feb 24 2020;doi:10.1016/j.ejpn.2020.02.011

101. Munch TN, Gortz S, Hauerberg J, Wohlfahrt J, Melbye M. Prognosis regarding shunt revision and mortality among hydrocephalus patients below the age of 2 years and the association to patient-related risk factors. Acta Neurochir (Wien). Mar 26 2020;doi:10.1007/s00701-020-04299-5

102. Ostrowski LE, Yin W, Patel M, et al. Restoring ciliary function to differentiated primary ciliary dyskinesia cells with a lentiviral vector. Gene Ther. Mar 2014;21(3):253–61. doi:10.1038/gt.2013.79

103. Conductier G, Brau F, Viola A, et al. Melanin-concentrating hormone regulates beat frequency of ependymal cilia and ventricular volume. Nat Neurosci. Jul 2013;16(7):845–7. doi:10.1038/nn.3401

104. Carter CS, Vogel TW, Zhang Q, et al. Abnormal development of NG2+PDGFR-alpha+ neural progenitor cells leads to neonatal hydrocephalus in a ciliopathy mouse model. Nat Med. Dec 2012;18(12):1797–804. doi:10.1038/nm.2996

