## Supplementary material for "The genetic background of hydrocephalus in a population-based cohort: implication of ciliary involvement"

#### Hydrocephalus-associated genes with high-impact variants

*ASTN2* (OMIM *612856): Two variants were identified in two heterozygote carriers; c.1816C>T; (p.Arg606Trp) and c.2491A>G; (p.Lys831Glu). *ASTN2* encodes astrotactin 2 and interact with *ASTN1* in the neuronal membrane, which forms adhesions between neurons and astroglia as a neuronal cell-surface antigen. By regulating its expression on the neuronal surface, astrotactin 2 mediates the formation and release of neuronal-glial adhesions during neuronal migration in fetal life[1], processes mediated by primary cilia. Astrotactin 2 furthermore plays a role in ASD and other neurodevelopmental disorders.[1,2] Lionel et al. describe hydrocephalus in three of 61 patients with rare CNV of interest, one of them with co-occurring psychiatric disorder and another with macrocephaly.[2]

*B3GALNT2* (OMIM *610194): A single variant c.824_825dupTT; (p.Ile276fs) was identified in a heterozygote carrier. The variant (c.822_823dup, p.Ile276Leufs*26) has previously been associated with the congenital muscular dystrophy-dystroglycanopathy Walker-Warburg syndrome affecting two brothers, one of whom also exhibited signs of ASD.[3] *B3GALNT2* encodes Beta-1,3-N-Acetylgalactosaminyltransferase 2 which is involved in the glycosylation of alpha-dystroglycan (α-DG). Reduced glycosylation impairs the interaction of α-DG with extracellular matrix elements,[4] which may disrupt the signalling functions of primary cilia, and lead to brain malformations.[5] Furthermore, severe syndromic hydrocephalus has been described as part of *B3GALNT2*-related diseases.[6]

*B3GLCT* (OMIM *610308): Two variants were identified in two heterozygote carriers; c.992T>C; (p.Leu331Ser) and c.1241A>G; (p.Tyr414Cys). *B3GLCT* encodes Beta-1,3-

glucosyltransferase which glucosylates a sub-set of thrombospondin-1 elements,[7] consequently interfering with Notch-, and Wnt/β-catenin signalling.[8] Autosomal recessive inheritance of Peter-plus syndrome (PPS) causing mutations in *B3GLCT* have been reported,[9] and hydrocephalus has been described in foetal PPS.[10] Furthermore, Mouse B3glct mutants developed highly penetrant hydrocephalus.[11] *B3GLCT* has not previously been associated with ASD.

*CELSR2* (OMIM *604265): We identified six high-impact variants in *CELSR2* in seven heterozygote carriers: c.1033C>T; (p.Arg345Cys), c.4508C>A; (p.Ser1503Tyr), c.613dupG; (p.Ala205fs), c.3110G>A; (p.Arg1037His), c.5657C>T; (p.Pro1886Leu), and c.6458G>A; (p.Arg2153Gln). *CELSR2* encodes Cadherin EGF LAG Seven-Pass G-Type Receptor 2 (previously called Epidermal Growth Factor-Like 2), a non-classic-type cadherin located at the plasma membrane, which is postulated to be involved in cell adhesion and receptor-ligand interactions. *CELSR2* has been associated impaired ciliogenesis[12] and planar cell polarity.[13] Lack of cadherins, Celsr2, and Celsr3 impairs ependymal ciliogenesis leading to defective CSF dynamics and fatal hydrocephalus.[14] *CELSR2* has not previously been directly associated with ASD.

*CENPF* (OMIM *600236): We identified a single high-impact variant in *CENPF* in a heterozygote carrier*:* c.8157delA; (p.Lys2719fs). *CENPF* is an established ciliary gene[15] and encodes Microtubule-binding centromere protein F, which regulate microtubule dynamics in the axoneme of the cilium.[16] Dysfunction leads to impairment of cell migration, focal adhesion dynamics, and primary cilia formation and in the brain it facilitates migration of glial progenitor nuclei to the ventricular surface, where they undergo mitosis.[16-18] *CENPF* has been associated with Stromme syndrome (microcephaly and ocular anomalies), which is considered a ciliopathy with a wide spectrum of phenotypes,[19, 20] including hydrocephalus.[21, 22] CENP-F dysfunction have been associated with impaired cortical neurogenesis.[23]

*COL4A1* (OMIM *120130): We identified three high-impact variants in *COL4A1* in four heterozygote carriers*:* c.2084C>A; (p.Pro695His), c.4742A>C; (p.Tyr1581Ser), and c.2447C>T; (p.Pro816Leu). *COL4A1* encodes the ubiquitously expressed Collagen type IV, alpha-1 chain protein, an integral part of basement membranes,[24] which plays an important role in angiogenesis.[25] Autosomal dominant inheritance of *COL4A1* mutations have previously been associated with tortuosity of retinal vessels, hereditary angiopathy with nephropathy and aneurysms, brain small vessel disease with or without ocular anomalies, porencephaly 1, and schizencephaly. Various missense mutations in Col4A1/2 lead to brain malformation and intracerebral haemorrhage in knockout-mice.[26] Additionally, ex-vacuo dilation of parts of the ventricular system due to porencephaly has been reported in both mouse and human mutation carriers.[27] Type IV collagen defects has been associated with polycystic kidney disease, which is considered a ciliary disease.[28] *COL4A1* has not been directly associated with autism spectrum disorder.

*CRTAP* (OMIM *605497): We identified a single high-impact variant in *CRTAP* in a heterozygote carrier*:* c.719T>C; (p.Phe240Ser). *CRTAP* encodes cartilage associated protein and has been associated with Cole-Carpenter syndrome, which is osteogenesis imperfecta featuring craniosynostosis and occasionally hydrocephalus.[29, 30] Together with *P3H1* and *PPIB*, the gene encode for the components of the endoplasmic reticulum (ER) complex responsible for the 3-hydroxylation of specific proline residues in type I collagen.[31] This results in abnormal collagen type I, which assembles in disorganized fibres.[32] *CRTAP* has to the best of our knowledge not been associated with ciliary function or ASD.

*DAG1* (OMIM *128239): We identified two high-impact variants in *DAG1* in two heterozygote carriers: c.1893_1896delGAAA; (p.Lys631fs) and c.1022C>T; (p.Thr341Ile). *DAG1* encodes dystrophin-associated glycoprotein 1, which forms part of the dystrophin-associated proteins complex (DAPC) consisting of dystroglycans, sarcoglycans, dystrobrevins and syntrophins. DAPC components are expressed and regulated during the neuronal or astrocytic differentiation of neural stem/progenitor cells,[33] a process regulated by primary cilia. Autosomal recessive inheritance of *DAG1* mutations have been associated with Muscular dystrophy dystroglycanopathy, and limb-girdle syndrome. Furthermore, *DAG1* has been associated with Walker-Warburg syndrome,[34, 35] which includes hydrocephalus as well as abnormally thick, flat cortex with irregular pebbled cortical-white matter border on MRI, hydrocephalus, scattered small periventricular heterotopia and subependymal haemorrhages and calcifications, z-shaped brainstem, occipital encephalocele, vermian agenesis, and an elongated and thick tectum.[34] Although rare, ASD has been described in clinical reports of dystroglycanopathies.[36]

*DNAH5* (OMIM *603335): We identified two high-impact variants in *DNAH5* in two heterozygote carriers: c.9911C>T; (p.Ser3304Leu) and c.7741A>G; (p.Lys2581Glu). *DNAH5* encodes Dynein, axonemal, heavy chain 5, which is important for structure and function of the motile ependymal cilia.[15, 37] Although frequently seen in patients without co-occurring hydrocephalus, autosomal recessive inheritance of *DNAH5* mutations have been associated with PCD, with and without situs inversus. Furthermore, the presence of hydrocephalus may be related to severity of PCD.[38, 39] *Dnah5* has been associated with obstructive hydrocephalus due to aqueductal stenosis in mouse models.[38] *DNAH5* has not previously been associated with ASD.

*DNAI1* (OMIM *604366): We identified a single high-impact variant in *DNAI1* in a heterozygote carrier*:* c.370C>T; (p.Arg124Cys). *DNAI1* encodes dynein in the outer dynein arms of the ciliary axoneme, intermediate chain 1 and mutations of this gene is associated with primary ciliary dyskinesia.[40] Lethal hydrocephalus is frequently seen in murine models of primary ciliary dyskinesia.[41] *DNAI1* or primary ciliary dyskinesia has not previously been associated with ASD.

*FLNA* (OMIM *300017): We identified two high-impact variants in *FLNA* in two homozygous carriers: c.2392G>A; (p.Glu798Lys) and c.3643G>A; (p.Gly1215Ser). *FLNA* encodes filamin A, alpha, an established ciliary gene,[15] which has a crucial role in ciliogenesis and basal body positioning. Additionally, the meckelin-filamin A signaling axis may be a key regulator in Wnt signalling.[42] X-linked dominant inheritance of *FLNA* mutations have been associated with periventricular heterotopia, Melnick-Needles syndrome, and X-linked recessive inheritance of *FLNA* mutations have been associated with FG Syndrome 2, Congenital short bowel syndrome, and Frontometaphyseal dysplasia 1, (OMIM *300017). *FLNA* associated with bilateral nodular periventricular heterotopia, also features foci of grey matter in abnormal locations in the brain secondary to arrest of neuronal migration as well as mega cisterna magna,[43] cerebellar hypoplasia, communicating hydrocephalus and bilateral hippocampal sclerosis.[44] *FLNA* mutations have been associated with non-syndromic ASD.[45]

*FREM1* (OMIM *608944): We identified two high-impact variants in *FREM1* in two heterozygote carriers*:* c.1493G>A; (p.Arg498Gln) and c.3874C>T; (p.Arg1292Cys). *FREM1* encodes FRAS1-related extracellular matrix protein 1, which plays a role in epidermal differentiation and epidermal adhesion during embryogenesis.[46] Additionally, in mice Fras1 is expressed in regions of the basement membrane that underlie organizing centres of the CNS, such as the roof plate of diencephalon, midbrain and hindbrain, as well as in the choroid plexus.[47] Autosomal dominant inheritance of *FREM1* mutations have been associated with trigonocephaly, (OMIN *608944). Furthermore, Severe hydrocephalus and shortened limbs have been described in a Chinese female foetus with *FREM1* mutations.[48] *FREM1* has not previously been associated with ASD.

*FUZ (OMIM* *610622)*:* We identified a single high-impact variant in *FUZ* in a heterozygote carrier*:* c.1211G>A; (p.Arg404Gln). *FUZ* encodes Fuzzy planar cell polarity protein. Fuz mutant mice display neural tube defects, skeletal dysmorphologies and Hedgehog signalling defects due to disrupted ciliogenesis.[49, 50] Planar cell polarity is of importance for the orientation of motile cilia and thereby also the directional cerebrospinal fluid flow and subsequent development of hydrocephalus.[51] Further, FUZ mutant cells lose functional primary cilia.[52] Autism spectrum disorders has not previously been linked with *FUZ* mutations.

*GBA* (OMIM *606463)*:* We identified a single high-impact variant in *GBA* in a heterozygote carrier*:* c.1279G>A; (p.Glu427Lys). *GBA* encodes beta-glucocerebrosidase, which breaks down glucosylceramide to glucose and ceramide within lysosomes.[53]Autosomal recessive inheritance of *GBA* mutations have been associated with the lysosomal storage disorder, Gaucher's disease.[53] *GBA* associated Gaucher's disease may also feature communicating hydrocephalus.[54-57] Interestingly, *GBA* mutations are numerically the greatest genetic risk factor for developing Parkinson's disease, the second most common neurodegenerative disorder.[58] *GBA* has not previously been associated with ASD.

*IFT172* (OMIM *607386)*:* We identified a single high-impact variant in *IFT172* in a heterozygote carrier*:* c.1036C>T; (p.Arg346*). *IFT172* is an established ciliary gene,[15] and encodes the intraflagellar transport protein 172, required for ciliogenesis and Hedgehog signalling.[59] Autosomal recessive inheritance of *IFT172I* mutations has been associated with retinitis pigmentosa, and short-rib dysplasia with or without polydactyly. In a mouse model, *Ift172* has been associated with the VACTERL-H (Vertebral, Anal, Cardiac, Tracheal-Esophageal, Renal, and Limb anomalies with hydrocephalus) phenotype.[59] *IFT172* has not been associated with ASD.

*KIAA1109* (OMIM *611565): We identified two high-impact variants in *KIAA1109* in two heterozygote carriers*:* c.14884A>G; (p.Thr4962Ala) and c.4927G>A; (p.Ala1643Thr). Homozygous or compound heterozygous mutations in *KIAA1109*, chromosome 4q27, is associated with Alkuraya-Kucinskas syndrome, according to OMIM, which may encompass congenital hydrocephalus and other brain malformations.[60, 61] *KIAA1109* has, to our knowledge, not previously been linked with ASD.

*LAMB1* (OMIM *150240): We identified a single high-impact variant in *LAMB1* in a heterozygote carrier: c.3199C>T; (p.Arg1067Cys). *LAMB1* encodes laminin, beta 1, an extracellular matrix protein involved in axonal outgrowth and guidance. Homozygous or compound heterozygous mutations in *LAMB1* has been associated with lissencephaly-V, which is an autosomal recessive brain malformation characterized by cobblestone changes in the cortex, more severe in the posterior region, and subcortical band heterotopia. This is a neuronal migration disorder characterized by protrusions of neurons beyond the first cortical layer at the pial surface of the brain, giving rise to the cobblestone changes. The cobblestone brain malformation is usually seen in association with congenital muscular dystrophies (dystroglycanopathies) and ocular abnormalities, also known as muscle-eye-brain disease. Affected individuals may have symptomatic hydrocephalus, porencephaly, seizures, and severely delayed psychomotor development.[62, 63] The role of *LAMB1* in ASD is uncertain, but *LAMB1* polymorphism has been associated with symptom severity in ASD in a single study.[64]

*LRP6* (OMIM *603507): We identified a single high-impact variant in *LRP6* in a heterozygote carrier: c.2203G>A; (p.Asp735Asn). *LRP6* encodes low density lipoprotein receptor-related protein 6, which play an important role as a co-receptor in Wnt/β-catenin signaling.[65] The binding of Lrp5/Lrp6 to the frizzled receptor initiates Wnt cascades.[65] Furthermore, Wnt pathway components, including Lrp6, regulate the centrosomal cycle and cilia formation and function.[66] An experimental study suggest involvement of Wnt/β-catenin signaling in hydrocephalus,[67] and a study of human foetuses with neural tube defect demonstrated involvement of the Wnt/β-catenin signaling component, DVL, in Dandy-Walker malformation.[68] *LRP6* has not previously been directly linked with ASD, but disruption of Wnt/β-catenin signaling has.[69]

*MPDZ* (OMIM *603785): We identified a single high-impact variant in *MPDZ* in a heterozygote carrier: c.18C>G; (p.Asp6Glu). *MPDZ* encodes the Multiple PDZ domain protein (MUPP1), which forms macromolecular complexes involved in signal transduction.[70] Autosomal recessive inheritance of *MPDZ* mutations has been associated with congenital hydrocephalus, with or without brain or eye anomalies, and compound heterozygotes have been associated with mild communicating hydrocephalus with enlarged subarachnoid space in a 9-month-old child, which resolved by the age of 17 months,[71] whereas humans and mice that carry a truncated version of MUPP1/Mupp1 display severe communicating hydrocephalus.[72] *Mpdz* loss-of-function mutations result in abnormally high CSF protein levels, as compared to normal mice.[72] Furthermore, the expression of the interacting planar cell polarity protein Pals1 is diminished in *Mpdz*-deficient mice, leading to progressive loss of the blood brain barrier integrity. The ependymal denudation was accompanied by reactive astrogliosis and subsequent aqueductal stenosis.[73] Contactin-associated protein-like 2 (CASPR2) is one of the synaptic adhesion molecules associated with ASD. CASPR2 forms a complex with receptors via interaction with MUPP1, which will lead to impaired CASPR2-MUPP1-GPR37 complex on the dendrites associated with one of the pathogeneses of ASD.[74]

*MTO1* (OMIM *614667): We identified a single high-impact variant in *MTO1* in a heterozygote carrier: c.369G>A; (p.Trp123*). *MTO1* encodes the mitochondrial translocation optimization 1 protein, which is involved in tRNA modification to increase the accuracy and efficiency of mtDNA translation.[75] Autosomal recessive inheritance of *MTO1* mutations has been associated with Combined oxidative phosphorylation deficiency 10, and Hypertrophic cardiomyopathy.[75] Neither hydrocephalus, nor ASD has been associated with mutations in *MTO1*, but the gene is not very well described. However, hydrocephalus has been associated with other gene mutations leading to mitochondrial dysfunction in mice, either because of impaired brain development or loss of ciliated epithelium in the ependymal layer.[76]

*NF1* (OMIM *613113): We identified two high-impact variants in *NF1* in two heterozygote carriers*:* c.1641+1G>C; (sp.) and c.5902C>T; (p.Arg1968*). *NF1* encodes neurofibromin 1, a tumour suppressor which is expressed largely in neuronal cells, it is known to associate with microtubules and is involved in several signaling pathways.[77] Autosomal dominant inheritance of *NF1* mutations has been associated to neurofibromatosis, and Watson syndrome. Although nervous system manifestations of neurofibromatosis is generally associated with optic nerve gliomas and neurofibromas, hydrocephalus is seen in up to 13 % of the patients.[78] In most cases, hydrocephalus is secondary to aqueductal stenosis or fourth ventricle webs,[79] others are caused by chiasmatic-hypothalamic tumours, and thalamic mass effect related to neurofibromatosis changes. [80] This process may link to the function of primary cilia, as these are required for the formation of neural stem cells located in the subventricular zones, primarily of the lateral ventricles in adults and the third ventricle and optic pathway in children as well as for neurogenesis through Hedgehog signalling.[81-83] High frequencies of early ASD and ADHD is seen among neurofibromatosis patients.[84, 85]

*NID1* (OMIM *131390): We identified a single high-impact variant in *NID1* in a heterozygote carrier: c.199G>T; (p.Asp67Tyr). *NID1* has according to OMIM been associated with autosomal dominant Dandy-Walker malformation and occipital cephalocele, which is a rare, congenital, and incompletely penetrant malformation that is considered part of the Dandy-Walker spectrum of disorders. Dandy-Walker malformation is a known cause of hydrocephalus. *NID1* encodes Nidogen 1 (previously called entactin), an extracellular matrix protein, which stabilizes the collagen IV and laminin networks of the blood brain barrier.[26] Primary cilia regulate extracellular matrix as part of their signaling functions.[86] Structural modelling of the *NID1-LAMC1* complex demonstrated that mutations in each of the genes disrupts the interaction.[87] Phenotype presentations include occipital cephalocele with a bony skull defect, and sometimes posterior fossa arachnoid cysts. Usually, the carriers display normal neurological development.[88] *NID1* has not previously been linked with ASD.

*NOTCH2* (OMIM *600275): We identified a single high-impact variant in *NOTCH2* in a heterozygote carrier: c.3430A>G; (p.Ser1144Gly). *NOTCH2* encodes the notch receptor 2 and is thereby essential for notch signalling and cortical neurogenesis, and has been linked with ASD and macocephaly.[89] Moreover, Notch2 and other Notch genes has been associated with choroid plexus tumours, a rare but distinct cause of hydrocephalus due to overproduction of CSF.[90, 91] Interestingly, primary cilia maintain homeostasis in mouse corneal epithelium cells through regulation of Notch signaling.[92]

*PIK3R2* (OMIM *603157): We identified a single high-impact variant in *PIK3R2* in a heterozygote carrier: c.1117G>A; (p.Gly373Arg). *PIKR3R2* encodes Phosphatidylinositol 3-kinase regulatory subunit 2, part of the PI3K-AKT-mTOR pathway, which is involved in angiogenesis, metabolism, cell proliferation and apoptosis.[93] *PIKR3R2* mutations have been associated with Megalencephaly-polymicrogyria-polydactyly-hydrocephalus syndrome (MPPH). MPPH syndrome is a developmental brain disorder characterized by oromotor dysfunction, epilepsy, intellectual disability and postaxial hexadactyly, enlarged brain size with bilateral perisylvian polymicrogyria and a variable degree of ventriculomegaly, around 50 % with hydrocephalus.[94, 95] PI3K-AKT-mTOR is related to megalencephaly syndromes encompassing intellectual disability and/or ASD.[96]

The particular *PIK3R2* variant, p.(Gly373Arg), is located in a regulatory subunit of the PI3K-AKT-mTOR pathway, and modulates GSK-3β activity, it was previously identified as a mosaic mutation and has been found to cause overgrowth syndromes.[97, 98] GSK-3β kinase is involved in the formation of centrosome-derived microtubules of the primary cilia and inhibition of GSK-3β results in reduced occurrence of primary cilia.[99]

*PLOD2* (OMIM *601865): We identified a single high-impact variant in *PLOD2* in a heterozygote carrier: c.695A>C; (p.Lys232Thr). *PLOD2* encodes Procollagen-lysine 2-oxyglutarate 5-dioxygenase 2, which is involved in the hydroxylation of collagen molecules.[100] Autosomal recessive inheritance of *PLOD2* mutations has been associated with Bruck syndrome. *PLOD2* has not been directly associated with hydrocephalus, but PLOD2 is expressed within the tunica media of the blood vessels in brain arterio-venous malformations (AVM's).[101] Both AVM and gliomas may cause hydrocephalus secondary to haemorrhage and obstruction, respectively. Dysregulation of *PLOD2* has furthermore been observed in gliomas; inhibition of *PLOD2* inactivated the PI3K/AKT signalling pathway and thus regulated the expression of its downstream epithelial-mesenchymal-transition-associated regulators, including E-cadherin, vimentin, N-cadherin, β-catenin, snail and slug in glioma cell.[101] ASD, together with skeletal abnormalities, has been associated with the *PLOD2* mutations.[102]

*POMT1* (OMIM *607423): We identified a single high-impact variant in *POMT1* in a heterozygote carrier: c.1216G>A; (p.Gly406Ser). *POMT1* encodes protein-O-mannosyltransferase 1, essential for alpha-dystroglycan (alpha-DG) glycosylation.[103] Autosomal recessive mutations in *POMT1* has been linked with severe congenital muscular dystrophy associated with defects in neuronal migration that produce complex brain and eye abnormalities, muscle-eye-brain disease (Walker-Warburg syndrome),[104] as well as cobblestone cortex (type II lissencephaly), hydrocephalus, agyria, thickened leptomeninges filled with neuroglial ectopia, disorganized cortical ribbon, and cerebellar dysplasia.[105] ASD has been described in the broad phenotypical spectrum of Walker-Warburg syndrome.[106]

*PTCH1* (OMIM *601309): We identified two high-impact variants in *PTCH* in two heterozygote carriers*:* c.1603-2A>C; (sp.) and c.3155C>T; (p.Thr1052Met). *PTCH1* encodes Patched 1, the Sonic hedgehog receptor.[107] Patched 1 an established ciliary gene important for neural tube development and regulation of ventricular zone neural stem cell fate.[108] Autosomal dominant inheritance of *PTCH1* mutations has been associated with Basal cell nevus syndrome (Gorlin syndrome) and holoprosencephaly. *PTCH1* has also been associated with obstructive hydrocephalus due to aqueductal stenosis in unrelated probands.[108] Furthermore, a disease related to Gorlin syndrome, the 9q22.3 microdeletion syndrome, presents with obstructive hydrocephalus, metopic craniosynostosis, overgrowth, developmental delay, intellectual disability, and seizures.[109] *PTCH1* has been associated with ASD in a case report of a female with

Gorlin syndrome as well as in a mouse model.[110, 111] The qk(v/v); ptch1$^{+/-}$ mice developed fatal acquired hydrocephalus due to aqueductal stenosis. The qk(v/v); ptch1$^{+/-}$ mice contained normal ciliated ependymal cells lining the ventricles of the brain, but these cells were functionally compromised with a severe cilia mediated flow defect.[112] Thus, *PTCH1* mutations may give rise to both dysfunction of the primary cilium as well as compromised ciliary motility. Interestingly, the ptch1 heterozygous mice were prone to several malignancies, including medulloblastomas, due to increased Sonic hedgehog signaling.[113]

*ROBO1:* We identified three high-impact variants in *ROBO1* in three heterozygote carriers: c.2665C>G; (p.Gln889Glu), c.979T>C; (p.Ser327Pro), and c.4683A>C; (p.Lys1561Asn). *ROBO1* encodes roundabout, axon guidance receptor, homolog 1, which is part of the immunoglobulin superfamily of cell adhesion molecules and expressed in CNS neurons.[114] Obstructive hydrocephalus due to stenosis of the Sylvian aqueduct has been described in Robo1/2 deficient mice in which Hes1 expression level was decreased in ventricular zone progenitors.[115] Hes1 is a major repressor of Notch signalling, and although Notch activation during early neural differentiation specifically promotes neural stem cells or early neural progenitors and delays their maturation, its inhibition promotes late neural progenitors and expedites neurogenesis, with a preference for neurons over glia. However, in combination with ciliary neurotrophic factor gliogenesis is promoted.[116] The involvement of Notch activation may link this proces with the function of primary cilia, which are necessary for the formation of neural stem cells and neurogenesis. *ROBO1* mutations has been associated with ASD in a population-based study from Faroe Islands.[117]

*SHOC2 (OMIM *602775):* We identified a single high-impact variant in *SHOC2* in a heterozygote carrier: c.1580A>G; (p.Asn527Ser). *SHOC2* encodes leucine-rich repeat scaffold protein, which is part of the RAS/MAPK pathway. Mutation SHOC2 and other genes in this pathway is associated with Noonan syndrome, which exhibit pre- and postnatal external hydrocephalus and sometimes Dandy-Walker malformation.[118, 119] To the best of our knowledge, SHOC2 has not yet been directly associated with ciliary function. Dysregulation of the RAS/MAPK signaling pathway is associated with ASD.[120]

*SLC12A3* (OMIM *600968): We identified a single high-impact variant in *SLC12A3* in a heterozygote carrier: c.2221G>A; (p.Gly741Arg) in a heterozygote carrier. *SLC12A3* encodes solute carrier family 12 (sodium/chloride transporters), member 3, and is mainly expressed in the kidneys and other organs, but not the brain. Homozygous and compound heterozygous mutations are associated with the recessively inheritied Gitelman syndrome, which have been associated with idiopathic intracranial hypertension (IIH). Thus, electrolyte abnormalities and secondary aldosteronism may be linked with IIH.[121, 122] A stillborn foetus with hydrocephalus of a mother with Gitelman syndrome has been reported.[123] To our knowledge, neither *SLC12A3* nor Gitelman syndrome has been linked with ASD.

*SMARCC1* (OMIM * 601732): We identified a single high-impact variant in *SMARCC1* in a heterozygote carrier: c.3224C>T; (p.Pro1075Leu). *SMARCC1* encodes SWI/SNF related, matrix associated, actin dependent regulator of chromatin, subfamily c, member 1, an ATP-dependent chromatin remodeler that regulates gene expression required for neural stem cell proliferation, differentiation, and survival during telencephalon development,[124] processes involving primary cilia-dependent signaling. *SMARCC1* has been associated with hydrocephalus due to aqueductal stenosis, as well as cardiac and skeletal abnormalities.[125] A study in zebrafish revealed interaction between Smarcc1a and the BBS6, which is involved in Bardet-Biedl syndrome, a well-described ciliopathy, of which 77% of cases display symptoms of ASD.[126]

*TRIM71* (OMIM. *618570): We identified a single high-impact variant in *TRIM71* in a heterozygote carrier: c.2432C>T; (p.Ala811Val) in a heterozygote carrier. *TRIM71* encodes tripartite motif-containing protein 71 and mutations cause communicating hydrocephalus with autosomal dominant inheritance. Furey et al. (2018) identified a de novo heterozygous missense mutations in *TRIM71* in three unrelated patients with congenital communicating hydrocephalus.[108] Jin et al. recently confirmed the finding of congenital communicating hydrocephalus in surgically treated patients with *TRIM71* mutations.[125] A mouse model shows that Lin41/Trim71 is essential for neurodevelopment and specifically is expressed in the ependymal cells lining the four brain ventricles which contains motile cilia.[127] To the best of our knowledge, *TRIM71* has not previously been linked with ASD.

*VANGL2:* We identified a single high-impact variant in *VANGL2* in a heterozygote carrier: c.1298C>T; (p.Thr433Met). *VANGL2* encodes VANGL planar cell polarity protein 2, which regulates polarity during development.[128] Autosomal dominant inheritance of *VANGL2* mutations has been associated with neural tube defects, including myelomeningocele with hydrocephalus,[13] and anencephaly.[129] *VANGL2* is an established ciliary gene, which plays a role in neural tube closure, planar cell polarity (PCP), and beating of motile ependymal cilia.[13, 15] Additionally, VANGL2 is involved in Wnt signalling.[130] The ciliary beating of multiciliated ependymal cells lining the walls of the brain ventricles are important for CSF flow. Key to this function is the rotational and translational PCP of the ependyma cells. Defects in the PCP can result in abnormal CSF accumulation and hydrocephalus.[131, 132] *VANGL2* has been associated with autism, possibly through interaction between Vangl2 and another PCP protein, the Prickle 2.[133]

*WDR81* (OMIM *614218): We identified a single high-impact variant in *WDR81* in a heterozygote carrier: c.796C>T; (p.Arg266Cys). WDR81 encodes WD repeat-containing domain 81 and mutation can cause recessively inherited congenital hydrocephalus as well as cerebellar ataxia, both with other brain malformations, according to OMIM. Recently, the human phenotype was expanded to foetal brain arrest and Dandy-Walker malformation.[134] WDR81 interacts with WDR91 (OMIM *616303) in an endosomal protein complex that inhibits PtdIns3 kinase, which is part of the PI3K-signaling pathway. To the best of our knowledge, *WDR81* has not been directly associated with ciliary function or ASD.

| Supplementary Table 1. Genes and transcripts screened | |
|---|---|
| **AKT3** | NM_001206729, NM_005465, NM_181690 |
| **ALG13** | NM_001039210, NM_001099922, NM_001168385, NM_001257230, NM_001257231, NM_001257234, NM_001257235, NM_001257237, NM_001257239, NM_001257240, NM_001257241, NM_001324290, NM_001324291, NM_001324292, NM_001324293, NM_001324294, NM_018466 |
| **AP1S2** | NM_001272071, NM_003916 |
| **ARFGEF2** | NM_006420 |
| **ARHGAP31** | NM_020754 |
| **ARSB** | NM_000046, NM_198709 |
| **ASTN2** | NM_001184734, NM_001184735, NM_014010, NM_198186, NM_198187, NM_198188 |
| **ATP6V0A2** | NM_012463 |
| **B3GALNT2** | NM_001277155, NM_152490 |
| **B3GLCT** | NM_194318 |
| **B4GAT1** | NM_006876 |
| **BMP4** | NM_001202, NM_001347912, NM_001347913, NM_001347914, NM_001347915, NM_001347916, NM_001347917, NM_130850, NM_130851 |
| **BRAF** | NM_004333 |
| **CBL** | NM_005188 |
| **CC2D2A** | NM_001080522, NM_001164720, NM_020785 |
| **CCDC88C** | NM_001080414 |
| **CCL2** | NM_002982 |
| **CCND2** | NM_001759 |
| **CELSR2** | NM_001408 |
| **CENPF** | NM_016343 |
| **CEP290** | NM_025114 |
| **CEP83** | NM_001042399, NM_001346457, NM_001346458, NM_001346459, NM_001346460, NM_001346461, NM_001346462, NM_016122 |
| **COL4A1** | NM_001303110, NM_001845 |
| **CRB2** | NM_173689 |
| **CREBBP** | NM_001079846, NM_004380 |
| **CRTAP** | NM_006371 |
| **DAG1** | NM_001165928, NM_001177634, NM_001177635, NM_001177636, NM_001177637, NM_001177638, NM_001177639, NM_001177640, NM_001177641, NM_001177642, NM_001177643, NM_001177644, NM_004393 |
| **DNAH5** | NM_001369 |
| **DNAI1** | NM_001281428, NM_012144 |
| **DOCK8** | NM_001190458, NM_001193536, NM_203447 |
| **DPH1** | NM_001346574, NM_001346575, NM_001346576, NM_001383 |
| **DYX1C1** | NM_001033559, NM_001033560, NM_130810 |
| **EML1** | NM_001008707, NM_004434 |
| **FANCB** | NM_001018113, NM_001324162, NM_152633 |
| **FGFR2** | NM_000141, NM_001144913, NM_001144914, NM_001144915, NM_001144916, NM_001144917, NM_001144918, NM_001144919, NM_001320654, NM_001320658, NM_022970, NM_023029 |
| **FGFR3** | NM_000142, NM_001163213, NM_022965 |
| **FKRP** | NM_001039885, NM_024301 |
| **FKTN** | NM_001079802, NM_001198963, NM_006731 |
| **FLNA** | NM_001110556, NM_001456 |
| **FMN2** | NM_020066.5, NM_001305424.2, NM_001348094.2 |
| **FOXJ1** | NM_001454.4 |

| | |
|---|---|
| **FOXP1** | NM_001012505, NM_001244808, NM_001244810, NM_001244812, NM_001244813, NM_001244814, NM_001244815, NM_001244816, NM_032682 |
| **FREM1** | NM_001177704, NM_144966 |
| **FUZ** | NM_001171937, NM_025129 |
| **FXYD2** | NM_001680.5, NM_021603.4 |
| **GBA** | NM_000157, NM_001005741, NM_001005742, NM_001171811, NM_001171812 |
| **Gene** | Transcripts |
| **GLI3** | NM_000168 |
| **GPSM2** | NM_001321038, NM_001321039, NM_013296 |
| **HIRA** | NM_003325 |
| **HRAS** | NM_001130442, NM_001318054, NM_005343, NM_176795 |
| **HYLS1** | NM_001134793, NM_145014 |
| **IFT172** | NM_015662 |
| **IGFBP4** | NM_001552 |
| **ISPD** | NM_001101417, NM_001101426 |
| **KCNG4** | NM_172347.2 |
| **KIAA0586** | NM_001244189, NM_001244190, NM_001244191, NM_001244192, NM_001244193, NM_001329943, NM_001329944, NM_001329945, NM_001329946, NM_001329947, NM_014749 |
| **KIAA1109** | NM_015312 |
| **KIF19** | NM_153209.4 |
| **KIF7** | NM_198525 |
| **KRAS** | NM_004985, NM_033360 |
| **L1CAM** | NM_000425, NM_001143963, NM_001278116, NM_024003 |
| **LAMB1** | NM_002291 |
| **LAMC1** | NM_002293 |
| **LARGE1** | NM_004737, NM_133642 |
| **LRP6** | NM_002336 |
| **MAF** | NM_001031804, NM_005360 |
| **MECP2** | NM_001110792, NM_001316337, NM_004992 |
| **MKS1** | NM_001165927, NM_001321268, NM_001321269, NM_001330397, NM_017777 |
| **MPDZ** | NM_001261406, NM_001261407, NM_001330637, NM_003829 |
| **MTM1** | NM_000252 |
| **MTO1** | NM_001123226, NM_012123, NM_133645 |
| **MTOR** | NM_004958.4, NM_001386500.1, NM_001386501.1 |
| **NF1** | NM_000267, NM_001042492, NM_001128147 |
| **NID1** | NM_002508 |
| **NLGN4X** | NM_001282145, NM_001282146, NM_020742, NM_181332 |
| **NOTCH2** | NM_001200001, NM_024408 |
| **NRAS** | NM_002524 |
| **OFD1** | NM_001330209, NM_001330210, NM_003611 |
| **P4HB** | NM_000918 |
| **PIK3CA** | NM_006218 |
| **PIK3R2** | NM_005027 |
| **PLOD2** | NM_000935, NM_182943 |
| **PMP22** | NM_000304, NM_001281455, NM_001281456, NM_001330143, NM_153321, NM_153322 |
| **POMGNT1** | NM_001243766, NM_001290129, NM_001290130, NM_017739 |

| | |
|---|---|
| **POMGNT2** | NM_032806 |
| **POMK** | NM_001277971, NM_032237 |
| **POMT1** | NM_001077365, NM_001077366, NM_001136113, NM_001136114, NM_007171 |
| **POMT2** | NM_013382 |
| **PROC** | NM_000312 |
| **PTCH1** | NM_000264, NM_001083602, NM_001083603, NM_001083604, NM_001083605, NM_001083606, NM_001083607 |
| **PTEN** | NM_000314.8, NM_001304717.5, NM_001304718.2 |
| **PTPN11** | NM_001330437, NM_002834, NM_080601 |
| **RAF1** | NM_002880 |
| **RHPN1** | NM_052924.3 |
| **ROBO1** | NM_001145845, NM_002941, NM_133631 |
| **RPS6KA3** | NM_004586 |
| **SHH** | NM_000193, NM_001310462 |
| **SHOC2** | NM_001269039, NM_001324336, NM_001324337, NM_007373 |
| **SIL1** | NM_001037633, NM_022464 |
| **SKI** | NM_003036 |
| **SLC12A3** | NM_000339, NM_001126107, NM_001126108 |
| **SLC9A6** | NM_001042537, NM_001177651, NM_001330652, NM_006359 |
| **SMARCC1** | NM_003074 |
| **SNX10** | NM_001199835, NM_001199837, NM_001199838, NM_001318198, NM_001318199, NM_013322 |
| **SOS1** | NM_005633 |
| **TBC1D7** | NM_001143964, NM_001143965, NM_001143966, NM_001258457, NM_001318805, NM_001318806, NM_016495 |
| **TBX1** | NM_005992, NM_080646, NM_080647 |
| **TBX15** | NM_001330677, NM_152380 |
| **TMEM216** | NM_001173990, NM_001173991, NM_001330285, NM_016499 |
| **TMEM5** | NM_001278237, NM_014254 |
| **TMEM67** | NM_001142301, NM_153704 |
| **TMEM92** | NM_001168215, NM_153229 |
| **TRIM71** | NM_001039111 |
| **TTR** | NM_000371 |
| **USP9X** | NM_001039590, NM_001039591 |
| **VANGL1** | NM_001172411, NM_001172412, NM_138959 |
| **VANGL2** | NM_020335 |
| **WDR81** | NM_001163809 |
| **ZIC1** | NM_003412 |
| **ZIC2** | NM_007129 |
| **ZIC3** | NM_001330661, NM_003413 |

**Supplementary Table 2. Indirect estimation of gene region coverage.**

| Symbol | Expected | Hit rate |
|---|---|---|
| **ALG13** | 8 | 50 |
| **ARFGEF2** | 6 | 100 |
| **ARHGAP31** | 7 | 85.7 |
| **ARSB** | 6 | 83.3 |
| **ASTN2** | 14 | 71.4 |
| **ATP6V0A2** | 8 | 87.5 |
| **B3GALNT2** | 2 | 50 |
| **B3GLCT** | 6 | 100 |
| **B4GAT1** | 2 | 50 |
| **BMP4** | 2 | 100 |
| **BRAF** | 2 | 100 |
| **CBL** | 1 | 100 |
| **CC2D2A** | 7 | 100 |
| **CCDC88C** | 20 | 85 |
| **CCL2** | 1 | 100 |
| **CCND2** | 1 | 100 |
| **CELSR2** | 27 | 88.9 |
| **CENPF** | 40 | 90 |
| **CEP290** | 14 | 57.1 |
| **CEP83** | 6 | 100 |
| **COL4A1** | 15 | 86.7 |
| **CRB2** | 24 | 45.8 |
| **CREBBP** | 20 | 90 |
| **CRPPA** | 4 | 75 |
| **CRTAP** | 6 | 100 |
| **DAG1** | 6 | 100 |
| **DNAAF4** | 4 | 75 |
| **DNAH5** | 35 | 97.1 |
| **DNAI1** | 2 | 100 |
| **DOCK8** | 25 | 92 |
| **DPH1** | 8 | 75 |
| **EML1** | 5 | 80 |
| **FANCB** | 1 | 100 |
| **FGFR2** | 5 | 100 |
| **FGFR3** | 12 | 83.3 |
| **FKRP** | 6 | 83.3 |
| **FKTN** | 6 | 100 |
| **FLNA** | 5 | 100 |
| **FMN2** | 60 | 20 |
| **FOXJ1** | 2 | 100 |
| **FOXP1** | 1 | 0 |
| **FREM1** | 26 | 96.2 |
| **FUZ** | 4 | 100 |
| **FXYD2** | 3 | 66.7 |

| | | |
|---|---|---|
| **GBA** | 2 | 100 |
| **GLI3** | 19 | 100 |
| **GPSM2** | 6 | 83.3 |
| **HIRA** | 1 | 100 |
| **HRAS** | 2 | 100 |
| **HYLS1** | 1 | 100 |
| **IFT172** | 6 | 100 |
| **IGFBP4** | 1 | 100 |
| **KCNG4** | 12 | 100 |
| **KIAA0586** | 12 | 75 |
| **KIAA1109** | 22 | 100 |
| **KIF19** | 15 | 100 |
| **KIF7** | 12 | 100 |
| **KRAS** | 1 | 0 |
| **L1CAM** | 2 | 100 |
| **LAMB1** | 13 | 100 |
| **LAMC1** | 19 | 89.5 |
| **LARGE1** | 6 | 100 |
| **LRP6** | 6 | 83.3 |
| **MAF** | 1 | 0 |
| **MECP2** | 5 | 80 |
| **MKS1** | 3 | 100 |
| **MPDZ** | 20 | 80 |
| **MTO1** | 5 | 80 |
| **MTOR** | 11 | 90.9 |
| **NF1** | 10 | 80 |
| **NID1** | 17 | 100 |
| **NLGN4X** | 4 | 50 |
| **NOTCH2** | 19 | 52.6 |
| **OFD1** | 1 | 100 |
| **P4HB** | 3 | 66.7 |
| **PIK3CA** | 3 | 66.7 |
| **PIK3R2** | 7 | 71.4 |
| **PLOD2** | 1 | 100 |
| **PMP22** | 1 | 100 |
| **POMGNT1** | 4 | 75 |
| **POMGNT2** | 2 | 100 |
| **POMK** | 1 | 100 |
| **POMT1** | 8 | 100 |
| **POMT2** | 3 | 100 |
| **PROC** | 2 | 100 |
| **PTCH1** | 9 | 100 |
| **RAF1** | 2 | 50 |
| **RHPN1** | 4 | 0 |
| **ROBO1** | 11 | 81.8 |
| **RPS6KA3** | 4 | 100 |
| **SHH** | 1 | 0 |

| | | |
|---|---|---|
| **SHOC2** | 1 | 100 |
| **SIL1** | 2 | 100 |
| **SKI** | 9 | 66.7 |
| **SLC12A3** | 12 | 100 |
| **SLC9A6** | 4 | 100 |
| **SMARCC1** | 1 | 100 |
| **SOS1** | 3 | 100 |
| **TBC1D7** | 2 | 100 |
| **TBX1** | 6 | 66.7 |
| **TBX15** | 4 | 100 |
| **TMEM216** | 3 | 66.7 |
| **TMEM67** | 6 | 83.3 |
| **TMEM92** | 3 | 33.3 |
| **TRIM71** | 3 | 66.7 |
| **TTR** | 2 | 100 |
| **USP9X** | 2 | 100 |
| **VANGL1** | 3 | 66.7 |
| **VANGL2** | 4 | 100 |
| **WDR81** | 13 | 23.1 |
| **ZIC1** | 1 | 100 |
| **ZIC2** | 5 | 60 |
| **ZIC3** | 2 | 50 |
| **NRAS** | 0 | non-expected |
| **AKT3** | 0 | non-expected |
| **SNX10** | 0 | non-expected |
| **RXYLT1** | 0 | non-expected |
| **PTPN11** | 0 | non-expected |
| **AP1S2** | 0 | non-expected |
| **MTM1** | 0 | non-expected |

**Supplementary Table 3. Identifying the best performing PHRED scores for discriminating high from low impact variants for each CADD consequence category.**

| Sequence ontology (CADD:Consequence) | PHRED-cutoff (High-impact) | Sensitivity | Specificity | F1-score | Method |
|---|---|---|---|---|---|
| **NON_SYNONYMOUS** | >23.25 | 0.712 | 0.886 | 0.756 | CART-algorithm |
| **INFRAME** | >18.99 | 0.811 | 0.807 | 0.736 | CART-algorithm |
| **SPLICE_SITE** | >18.18 | 0.975 | 0.590 | 0.685 | CART-algorithm |
| **FRAME_SHIFT** | - | 1.000 | 0.00 | 0.998 | Class = High-impact |
| **CANONICAL_SPLICE** | - | 1.000 | 0.00 | 0.994 | Class = High-impact |
| **STOP_GAINED** | - | 1.000 | 0.00 | 0.998 | Class = High-impact |
| **STOP_LOST** | - | 1.000 | 0.00 | 0.947 | Class = High-impact |

**Supplementary Table 4: Sensitivity analysis method and results analysing 121 genes in 72 hydrocephalus cases and 4,181 background population controls.**

| Variable | Sequence ontology (CADD:Consequence) | Range in percentage difference in allelic load (Cases-Controls) | P value range |
|---|---|---|---|
| **AF** | non-synonomous | 0.0028 - 0.0102 | 0.036 - 1 |
| **AF** | in-frame* | -0.0119 - -0.0518 | 0.471 - 1 |
| **AF** | splice-site* | -0.0167 - -0.0169 | 1.0 - 1.0 |
| **AF** | PTV’s | 0.0276 - 0.0308 | 0.002 - 0.008 |
| **PHRED** | non_synonomous | -0.0031-0.0053 | 0.161 - 1 |

*Only identified in the control population
AF: allele frequency.

**Supplementary Table 5. Sensitivity analysis for High-impact, multi-hit results.**

| Variable: | # Hits | % diff. Range (Cases-Controls) | P value range |
|---|---|---|---|
| **AF - PHRED** | 1 | -2.32 -17.33 | 0.003 -1 |
| **AF - PHRED** | 2 | -10.78 - 5.96 | 0.033 - 1 |
| **AF - PHRED** | 3 | -2.87 - 10.75 | 0.009 - 1 |
| **AF - PHRED** | 4 | -1.412 - 5.63 | 0.030 - 1 |
| **AF - PHRED** | 5 | -1.89 - 4.66 | 0.026 - 1 |

**Supplementary Table 6. Distribution of high-impact variants in the subset of 34 hydrocephalus candidate genes found in 72 hydrocephalus patients and in 4,181 background population controls.**

| | Hydrocephalus (N=72) | Proportion of cases (%) | Controls (N=4,181) | Proportion of controls (%) |
|---|---|---|---|---|
| **Number of genes** | 34 | | | |
| **High-impact variants identified** | 52 | - | 810 | - |
| **High-impact variant carriers** | 42 | 58.3 | 1,210 | 28.9 |
| carriers with 2 + variants | 10 | 13.9 | 215 | 5.1 |
| carriers with 1 variant | 32 | 44.4 | 995 | 23.8 |
| non-carriers (0 variants) | 30 | 41.7 | 2,971 | 71.1 |
| **Homozygote High-impact variant carriers** | 0 | 0 | 1 | 0.02 |
| **Hemizygote High-impact variant carriers** | 2 | 2.78 | 4 | 0.1 |

**Supplementary Table 7. Types of high-impact variants in the subset of 34 hydrocephalus candidate genes and in 4,181 background population controls.**

| | Hydrocephalus cases (N=72) | Proportion of high impact variants (%) | Controls (N=4,181) | Proportion of high impact variants (%) |
|---|---|---|---|---|
| **Protein truncating variants** | 9 | 17.3 | 64 | 7.9 |
| **canonical splice** | 2 | 3.9 | 18 | 2.2 |
| **frame-shift** | 4 | 7.7 | 23 | 2.8 |
| **stop gained** | 3 | 5.8 | 23 | 2.8 |
| **stop lost** | 0 | 0 | 0 | 0 |
| **Non-protein truncating variants** | 43 | 82.7 | 746 | 92.1 |
| **non-synonymous** | 43 | 82.7 | 737 | 90.9 |
| **splice-site** | 0 | 0 | 6 | 0.74 |
| **in-frame** | 0 | 0 | 3 | 0.37 |

### References


1. Wilson PM, Fryer RH, Fang Y, Hatten ME. Astn2, a novel member of the astrotactin gene family, regulates the trafficking of ASTN1 during glial-guided neuronal migration. *J Neurosci*. Jun 23 2010;30(25):8529-40. doi:10.1523/jneurosci.0032-10.2010
2. Lionel AC, Tammimies K, Vaags AK, et al. Disruption of the ASTN2/TRIM32 locus at 9q33.1 is a risk factor in males for autism spectrum disorders, ADHD and other neurodevelopmental phenotypes. *Hum Mol Genet*. May 15 2014;23(10):2752-68. doi:10.1093/hmg/ddt669
3. Maroofian R, Riemersma M, Jae LT, et al. B3GALNT2 mutations associated with non-syndromic autosomal recessive intellectual disability reveal a lack of genotype-phenotype associations in the muscular dystrophy-dystroglycanopathies. *Genome Med*. Dec 22 2017;9(1):118. doi:10.1186/s13073-017-0505-2
4. Stevens E, Carss KJ, Cirak S, et al. Mutations in B3GALNT2 cause congenital muscular dystrophy and hypoglycosylation of alpha-dystroglycan. *Am J Hum Genet*. Mar 7 2013;92(3):354-65. doi:10.1016/j.ajhg.2013.01.016
5. Nickolls AR, Lee MM, Zukosky K, Mallon BS, Bönnemann CG. Human embryoid bodies as a 3D tissue model of the extracellular matrix and α-dystroglycanopathies. *Dis Model Mech*. Jun 26 2020;13(6)doi:10.1242/dmm.042986
6. Al Dhaibani MA, El-Hattab AW, Ismayl O, Suleiman J. B3GALNT2-Related Dystroglycanopathy: Expansion of the Phenotype with Novel Mutation Associated with Muscle-Eye-Brain Disease, Walker-Warburg Syndrome, Epileptic Encephalopathy-West Syndrome, and Sensorineural Hearing Loss. *Neuropediatrics*. Aug 2018;49(4):289-295. doi:10.1055/s-0038-1651519
7. Du J, Takeuchi H, Leonhard-Melief C, et al. O-fucosylation of thrombospondin type 1 repeats restricts epithelial to mesenchymal transition (EMT) and maintains epiblast pluripotency during mouse gastrulation. *Dev Biol*. Oct 1 2010;346(1):25-38. doi:10.1016/j.ydbio.2010.07.008
8. Vasudevan D, Takeuchi H, Johar SS, Majerus E, Haltiwanger RS. Peters plus syndrome mutations disrupt a noncanonical ER quality-control mechanism. *Curr Biol*. Feb 2 2015;25(3):286-295. doi:10.1016/j.cub.2014.11.049
9. Reis LM, Tyler RC, Abdul-Rahman O, et al. Mutation analysis of B3GALTL in Peters Plus syndrome. *Am J Med Genet A*. Oct 15 2008;146A(20):2603-10. doi:10.1002/ajmg.a.32498
10. Schoner K, Kohlhase J, Müller AM, et al. Hydrocephalus, agenesis of the corpus callosum, and cleft lip/palate represent frequent associations in fetuses with Peters' plus syndrome and B3GALTL mutations. Fetal PPS phenotypes, expanded by Dandy Walker cyst and encephalocele. *Prenat Diagn*. Jan 2013;33(1):75-80. doi:10.1002/pd.4012
11. Holdener BC, Percival CJ, Grady RC, et al. ADAMTS9 and ADAMTS20 are differentially affected by loss of B3GLCT in mouse model of Peters plus syndrome. *Hum Mol Genet*. Dec 15 2019;28(24):4053-4066. doi:10.1093/hmg/ddz225
12. Shaheen R, Szymanska K, Basu B, et al. Characterizing the morbid genome of ciliopathies. *Genome Biol*. Nov 28 2016;17(1):242. doi:10.1186/s13059-016-1099-5

13. Yamasaki M, Kanemura Y. Molecular Biology of Pediatric Hydrocephalus and Hydrocephalus-related Diseases. *Neurol Med Chir (Tokyo)*. 2015;55(8):640-6. doi:10.2176/nmc.ra.2015-0075
14. Tissir F, Qu Y, Montcouquiol M, et al. Lack of cadherins Celsr2 and Celsr3 impairs ependymal ciliogenesis, leading to fatal hydrocephalus. *Nat Neurosci*. Jun 2010;13(6):700-7. doi:10.1038/nn.2555
15. van Dam TJP, Kennedy J, van der Lee R, et al. CiliaCarta: An integrated and validated compendium of ciliary genes. *PLoS One*. 2019;14(5):e0216705. doi:10.1371/journal.pone.0216705
16. Pfaltzgraff ER, Roth GM, Miller PM, Gintzig AG, Ohi R, Bader DM. Loss of CENP-F results in distinct microtubule-related defects without chromosomal abnormalities. *Mol Biol Cell*. Jul 1 2016;27(13):1990-9. doi:10.1091/mbc.E15-12-0848
17. Hu DJ, Baffet AD, Nayak T, Akhmanova A, Doye V, Vallee RB. Dynein recruitment to nuclear pores activates apical nuclear migration and mitotic entry in brain progenitor cells. *Cell*. Sep 12 2013;154(6):1300-13. doi:10.1016/j.cell.2013.08.024
18. Baffet AD, Hu DJ, Vallee RB. Cdk1 Activates Pre-mitotic Nuclear Envelope Dynein Recruitment and Apical Nuclear Migration in Neural Stem Cells. *Dev Cell*. Jun 22 2015;33(6):703-16. doi:10.1016/j.devcel.2015.04.022
19. Ozkinay F, Atik T, Isik E, et al. A further family of Stromme syndrome carrying CENPF mutation. *Am J Med Genet A*. Jun 2017;173(6):1668-1672. doi:10.1002/ajmg.a.38173
20. Filges I, Bruder E, Brandal K, et al. Strømme Syndrome Is a Ciliary Disorder Caused by Mutations in CENPF. *Hum Mutat*. Apr 2016;37(4):359-63. doi:10.1002/humu.22960
21. Slee J, Goldblatt J. Further evidence for a syndrome of "apple peel" intestinal atresia, ocular anomalies and microcephaly. *Clin Genet*. Oct 1996;50(4):260-2. doi:10.1111/j.1399-0004.1996.tb02640.x
22. Alghamdi M, Alkhamis WH, Bashiri FA, et al. Expanding the phenotype and the genotype of Stromme syndrome: A novel variant of the CENPF gene and literature review. *Eur J Med Genet*. May 2020;63(5):103844. doi:10.1016/j.ejmg.2020.103844
23. Waters AM, Asfahani R, Carroll P, et al. The kinetochore protein, CENPF, is mutated in human ciliopathy and microcephaly phenotypes. *J Med Genet*. Mar 2015;52(3):147-56. doi:10.1136/jmedgenet-2014-102691
24. Timpl R, Aumailley M. Biochemistry of basement membranes. *Adv Nephrol Necker Hosp*. 1989;18:59-76.
25. Bahramsoltani M, Slosarek I, De Spiegelaere W, Plendl J. Angiogenesis and collagen type IV expression in different endothelial cell culture systems. *Anat Histol Embryol*. Apr 2014;43(2):103-15. doi:10.1111/ahe.12052
26. Xu L, Nirwane A, Yao Y. Basement membrane and blood-brain barrier. *Stroke Vasc Neurol*. Jul 2019;4(2):78-82. doi:10.1136/svn-2018-000198
27. Verbeek E, Meuwissen ME, Verheijen FW, et al. COL4A2 mutation associated with familial porencephaly and small-vessel disease. *Eur J Hum Genet*. Aug 2012;20(8):844-51. doi:10.1038/ejhg.2012.20
28. Cornec-Le Gall E, Chebib FT, Madsen CD, et al. The Value of Genetic Testing in Polycystic Kidney Diseases Illustrated by a Family With PKD2 and COL4A1 Mutations. *Am J Kidney Dis*. Aug 2018;72(2):302-308. doi:10.1053/j.ajkd.2017.11.015

29. Balasubramanian M, Pollitt RC, Chandler KE, et al. CRTAP mutation in a patient with Cole-Carpenter syndrome. *Am J Med Genet A*. Mar 2015;167a(3):587-91. doi:10.1002/ajmg.a.36916
30. MacDermot KD, Buckley B, Van Someren V. Osteopenia, abnormal dentition, hydrops fetalis and communicating hydrocephalus. *Clin Genet*. Oct 1995;48(4):217-20. doi:10.1111/j.1399-0004.1995.tb04092.x
31. Besio R, Garibaldi N, Leoni L, et al. Cellular stress due to impairment of collagen prolyl hydroxylation complex is rescued by the chaperone 4-phenylbutyrate. *Dis Model Mech*. Jun 20 2019;12(6)doi:10.1242/dmm.038521
32. Tonelli F, Cotti S, Leoni L, et al. Crtap and p3h1 knock out zebrafish support defective collagen chaperoning as the cause of their osteogenesis imperfecta phenotype. *Matrix Biol*. Aug 2020;90:40-60. doi:10.1016/j.matbio.2020.03.004
33. Romo-Yáñez J, Rodríguez-Martínez G, Aragón J, et al. Characterization of the expression of dystrophins and dystrophin-associated proteins during embryonic neural stem/progenitor cell differentiation. *Neurosci Lett*. Sep 25 2020;736:135247. doi:10.1016/j.neulet.2020.135247
34. Leibovitz Z, Mandel H, Falik-Zaccai TC, et al. Walker-Warburg syndrome and tectocerebellar dysraphia: A novel association caused by a homozygous DAG1 mutation. *Eur J Paediatr Neurol*. May 2018;22(3):525-531. doi:10.1016/j.ejpn.2017.12.012
35. Riemersma M, Mandel H, van Beusekom E, et al. Absence of α- and β-dystroglycan is associated with Walker-Warburg syndrome. *Neurology*. May 26 2015;84(21):2177-82. doi:10.1212/wnl.0000000000001615
36. Astrea G, Romano A, Angelini C, et al. Broad phenotypic spectrum and genotype-phenotype correlations in GMPPB-related dystroglycanopathies: an Italian cross-sectional study. *Orphanet J Rare Dis*. Sep 26 2018;13(1):170. doi:10.1186/s13023-018-0863-x
37. Roberts AJ, Kon T, Knight PJ, Sutoh K, Burgess SA. Functions and mechanics of dynein motor proteins. *Nat Rev Mol Cell Biol*. Nov 2013;14(11):713-26. doi:10.1038/nrm3667
38. Ibañez-Tallon I, Gorokhova S, Heintz N. Loss of function of axonemal dynein Mdnah5 causes primary ciliary dyskinesia and hydrocephalus. *Hum Mol Genet*. Mar 15 2002;11(6):715-21. doi:10.1093/hmg/11.6.715
39. Ibanez-Tallon I, Pagenstecher A, Fliegauf M, et al. Dysfunction of axonemal dynein heavy chain Mdnah5 inhibits ependymal flow and reveals a novel mechanism for hydrocephalus formation. *Hum Mol Genet*. Sep 15 2004;13(18):2133-41. doi:10.1093/hmg/ddh219
40. Ostrowski LE, Yin W, Rogers TD, et al. Conditional deletion of dnaic1 in a murine model of primary ciliary dyskinesia causes chronic rhinosinusitis. *Am J Respir Cell Mol Biol*. Jul 2010;43(1):55-63. doi:10.1165/rcmb.2009-0118OC
41. Cho KJ, Noh SH, Han SM, et al. ZMYND10 stabilizes intermediate chain proteins in the cytoplasmic pre-assembly of dynein arms. *PLoS Genet*. Mar 2018;14(3):e1007316. doi:10.1371/journal.pgen.1007316
42. Adams M, Simms RJ, Abdelhamed Z, et al. A meckelin-filamin A interaction mediates ciliogenesis. *Hum Mol Genet*. Mar 15 2012;21(6):1272-86. doi:10.1093/hmg/ddr557
43. Parrini E, Ramazzotti A, Dobyns WB, et al. Periventricular heterotopia: phenotypic heterogeneity and correlation with Filamin A mutations. *Brain*. Jul 2006;129(Pt 7):1892-906. doi:10.1093/brain/awl125

44. Gurusamy S, Saravanan S, Kuttuva Premnath U, Sivanandan R. Bilateral posterior periventricular nodular heterotopia with cerebellar hypoplasia, communicating hydrocephalus and bilateral hippocampal sclerosis. A case report. *Neuroradiol J*. Mar 23 2009;22(1):11-5. doi:10.1177/197140090902200102
45. Sakai Y, Shaw CA, Dawson BC, et al. Protein interactome reveals converging molecular pathways among autism disorders. *Sci Transl Med*. Jun 8 2011;3(86):86ra49. doi:10.1126/scitranslmed.3002166
46. Smyth I, Du X, Taylor MS, Justice MJ, Beutler B, Jackson IJ. The extracellular matrix gene Frem1 is essential for the normal adhesion of the embryonic epidermis. *Proc Natl Acad Sci U S A*. Sep 14 2004;101(37):13560-5. doi:10.1073/pnas.0402760101
47. Makrygiannis AK, Pavlakis E, Petrou P, Kalogeraki E, Chalepakis G. Segmental and restricted localization pattern of Fras1 in the developing meningeal basement membrane in mouse. *Histochem Cell Biol*. Nov 2013;140(5):595-601. doi:10.1007/s00418-013-1150-5
48. Yang YD, Huang LY, Yan JM, Han J, Zhang Y, Li DZ. Novel FREM1 mutations are associated with severe hydrocephalus and shortened limbs in a prenatal case. *Eur J Obstet Gynecol Reprod Biol*. Aug 2017;215:262-264. doi:10.1016/j.ejogrb.2017.06.017
49. Gray RS, Abitua PB, Wlodarczyk BJ, et al. The planar cell polarity effector Fuz is essential for targeted membrane trafficking, ciliogenesis and mouse embryonic development. *Nat Cell Biol*. Oct 2009;11(10):1225-32. doi:10.1038/ncb1966
50. Zhang W, Taylor SP, Ennis HA, et al. Expanding the genetic architecture and phenotypic spectrum in the skeletal ciliopathies. *Hum Mutat*. Jan 2018;39(1):152-166. doi:10.1002/humu.23362
51. Takagishi M, Sawada M, Ohata S, et al. Daple Coordinates Planar Polarized Microtubule Dynamics in Ependymal Cells and Contributes to Hydrocephalus. *Cell Rep*. Jul 25 2017;20(4):960-972. doi:10.1016/j.celrep.2017.06.089
52. Barrell WB, Adel Al-Lami H, Goos JAC, et al. Identification of a novel variant of the ciliopathic gene FUZZY associated with craniosynostosis. *Eur J Hum Genet*. Nov 1 2021;doi:10.1038/s41431-021-00988-6
53. Boer DEC, van Smeden J, Bouwstra JA, Aerts J. Glucocerebrosidase: Functions in and Beyond the Lysosome. *J Clin Med*. Mar 9 2020;9(3)doi:10.3390/jcm9030736
54. Stone DL, Tayebi N, Coble C, Ginns EI, Sidransky E. Cardiovascular fibrosis, hydrocephalus, ophthalmoplegia, and visceral involvement in an American child with Gaucher disease. *J Med Genet*. Nov 2000;37(11):E40. doi:10.1136/jmg.37.11.e40
55. Cindik N, Ozcay F, Süren D, et al. Gaucher disease with communicating hydrocephalus and cardiac involvement. *Clin Cardiol*. Jan 2010;33(1):E26-30. doi:10.1002/clc.20348
56. Shiihara T, Oka A, Suzaki I, Ida H, Takeshita K. Communicating hydrocephalus in a patient with Gaucher's disease type 3. *Pediatr Neurol*. Mar 2000;22(3):234-6. doi:10.1016/s0887-8994(99)00140-x
57. Erduran E, Mocan H, Gedik Y, Kamaci R, Okten A, Değer O. Hydrocephalus, corneal opacities, deafness, left ventricle hypertrophy, clinodactyly in an adolescent patient. A new syndrome associated with glucocerebrosidase deficiency. *Genet Couns*. 1995;6(3):211-5.

58. Gegg ME, Schapira AH. Mitochondrial dysfunction associated with glucocerebrosidase deficiency. *Neurobiol Dis*. Jun 2016;90:43-50. doi:10.1016/j.nbd.2015.09.006
59. Friedland-Little JM, Hoffmann AD, Ocbina PJ, et al. A novel murine allele of Intraflagellar Transport Protein 172 causes a syndrome including VACTERL-like features with hydrocephalus. *Hum Mol Genet*. Oct 1 2011;20(19):3725-37. doi:10.1093/hmg/ddr241
60. Kumar K, Bellad A, Prasad P, Girimaji SC, Muthusamy B. KIAA1109 gene mutation in surviving patients with Alkuraya-Kučinskas syndrome: a review of literature. *BMC Med Genet*. Jun 26 2020;21(1):136. doi:10.1186/s12881-020-01074-2
61. Meszarosova AU, Lastuvkova J, Rennerova L, et al. Two novel pathogenic variants in KIAA1109 causing Alkuraya-Kučinskas syndrome in two Czech Roma brothers. *Clin Dysmorphol*. Oct 2020;29(4):197-201. doi:10.1097/mcd.0000000000000335
62. Radmanesh F, Caglayan AO, Silhavy JL, et al. Mutations in LAMB1 cause cobblestone brain malformation without muscular or ocular abnormalities. *Am J Hum Genet*. Mar 7 2013;92(3):468-74. doi:10.1016/j.ajhg.2013.02.005
63. Tonduti D, Dorboz I, Renaldo F, et al. Cystic leukoencephalopathy with cortical dysplasia related to LAMB1 mutations. *Neurology*. May 26 2015;84(21):2195-7. doi:10.1212/wnl.0000000000001607
64. Kim YJ, Park JK, Kang WS, et al. LAMB1 polymorphism is associated with autism symptom severity in Korean autism spectrum disorder patients. *Nord J Psychiatry*. 2015;69(8):594-8. doi:10.3109/08039488.2015.1022597
65. Bryja V, Červenka I, Čajánek L. The connections of Wnt pathway components with cell cycle and centrosome: side effects or a hidden logic? *Crit Rev Biochem Mol Biol*. Dec 2017;52(6):614-637. doi:10.1080/10409238.2017.1350135
66. Gupta GD, Coyaud É, Gonçalves J, et al. A Dynamic Protein Interaction Landscape of the Human Centrosome-Cilium Interface. *Cell*. Dec 3 2015;163(6):1484-99. doi:10.1016/j.cell.2015.10.065
67. Xu H, Xu B, Wang Z, Tan G, Shen S. Inhibition of Wnt/β-catenin signal is alleviated reactive gliosis in rats with hydrocephalus. *Childs Nerv Syst*. Feb 2015;31(2):227-34. doi:10.1007/s00381-014-2613-2
68. Liu L, Liu W, Shi Y, et al. DVL mutations identified from human neural tube defects and Dandy-Walker malformation obstruct the Wnt signaling pathway. *J Genet Genomics*. Jun 20 2020;47(6):301-310. doi:10.1016/j.jgg.2020.06.003
69. Alexander JM, Pirone A, Jacob MH. Excessive β-Catenin in Excitatory Neurons Results in Reduced Social and Increased Repetitive Behaviors and Altered Expression of Multiple Genes Linked to Human Autism. *Front Synaptic Neurosci*. 2020;12:14. doi:10.3389/fnsyn.2020.00014
70. Ullmer C, Schmuck K, Figge A, Lübbert H. Cloning and characterization of MUPP1, a novel PDZ domain protein. *FEBS Lett*. Mar 6 1998;424(1-2):63-8. doi:10.1016/s0014-5793(98)00141-0
71. Al-Jezawi NK, Al-Shamsi AM, Suleiman J, et al. Compound heterozygous variants in the multiple PDZ domain protein (MPDZ) cause a case of mild non-progressive communicating hydrocephalus. *BMC Med Genet*. Mar 2 2018;19(1):34. doi:10.1186/s12881-018-0540-x
72. Yang J, Simonneau C, Kilker R, et al. Murine MPDZ-linked hydrocephalus is caused by hyperpermeability of the choroid plexus. *EMBO Mol Med*. Jan 2019;11(1)doi:10.15252/emmm.201809540

73. Feldner A, Adam MG, Tetzlaff F, et al. Loss of Mpdz impairs ependymal cell integrity leading to perinatal-onset hydrocephalus in mice. *EMBO Mol Med*. Jul 2017;9(7):890-905. doi:10.15252/emmm.201606430
74. Tanabe Y, Fujita-Jimbo E, Momoi MY, Momoi T. CASPR2 forms a complex with GPR37 via MUPP1 but not with GPR37(R558Q), an autism spectrum disorder-related mutation. *J Neurochem*. Aug 2015;134(4):783-93. doi:10.1111/jnc.13168
75. Ghezzi D, Baruffini E, Haack TB, et al. Mutations of the mitochondrial-tRNA modifier MTO1 cause hypertrophic cardiomyopathy and lactic acidosis. *Am J Hum Genet*. Jun 8 2012;90(6):1079-87. doi:10.1016/j.ajhg.2012.04.011
76. Delavallée L, Mathiah N, Cabon L, et al. Mitochondrial AIF loss causes metabolic reprogramming, caspase-independent cell death blockade, embryonic lethality, and perinatal hydrocephalus. *Mol Metab*. May 30 2020;40:101027. doi:10.1016/j.molmet.2020.101027
77. Trovó-Marqui AB, Tajara EH. Neurofibromin: a general outlook. *Clin Genet*. Jul 2006;70(1):1-13. doi:10.1111/j.1399-0004.2006.00639.x
78. Roth J, Constantini S, Cinalli G. Neurofibromatosis type 1-related hydrocephalus: causes and treatment considerations. *Childs Nerv Syst*. Jun 6 2020;doi:10.1007/s00381-020-04719-y
79. Murphy C, Vassallo G, Burkitt-Wright E, et al. A retrospective regional study of aqueduct stenosis and fourth ventricle outflow obstruction in the paediatric complex neurofibromatosis type 1 population; Aetiology, clinical presentation and management. *Clin Neurol Neurosurg*. Jun 2020;193:105791. doi:10.1016/j.clineuro.2020.105791
80. Dincer A, Yener U, Ozek MM. Hydrocephalus in patients with neurofibromatosis type 1: MR imaging findings and the outcome of endoscopic third ventriculostomy. *AJNR Am J Neuroradiol*. Apr 2011;32(4):643-6. doi:10.3174/ajnr.A2357
81. Dahiya S, Lee DY, Gutmann DH. Comparative characterization of the human and mouse third ventricle germinal zones. *J Neuropathol Exp Neurol*. Jul 2011;70(7):622-33. doi:10.1097/NEN.0b013e31822200aa
82. Han YG, Spassky N, Romaguera-Ros M, et al. Hedgehog signaling and primary cilia are required for the formation of adult neural stem cells. *Nat Neurosci*. Mar 2008;11(3):277-84. doi:10.1038/nn2059
83. Tong CK, Han YG, Shah JK, Obernier K, Guinto CD, Alvarez-Buylla A. Primary cilia are required in a unique subpopulation of neural progenitors. *Proc Natl Acad Sci U S A*. Aug 26 2014;111(34):12438-43. doi:10.1073/pnas.1321425111
84. Kolesnik AM, Jones EJH, Garg S, Green J, Charman T, Johnson MH. Early development of infants with neurofibromatosis type 1: a case series. *Mol Autism*. 2017;8:62. doi:10.1186/s13229-017-0178-0
85. Philip R, Turk J. Neurofibromatosis and Attentional Deficits: An Illustrative Example of the Common Association of Medical Causes with Behavioural Syndromes, Implications for General Child Mental Health Services. *Child Adolesc Ment Health*. May 2006;11(2):89-93. doi:10.1111/j.1475-3588.2006.00398.x
86. Collins I, Wann AKT. Regulation of the Extracellular Matrix by Ciliary Machinery. *Cells*. Jan 23 2020;9(2)doi:10.3390/cells9020278
87. Darbro BW, Mahajan VB, Gakhar L, et al. Mutations in extracellular matrix genes NID1 and LAMC1 cause autosomal dominant Dandy-Walker malformation and occipital cephaloceles. *Hum Mutat*. Aug 2013;34(8):1075-9. doi:10.1002/humu.22351

88. McNiven V, Ito YA, Hartley T, Kernohan K, Miller E, Armour CM. NID1 variant associated with occipital cephaloceles in a family expressing a spectrum of phenotypes. *Am J Med Genet A*. May 2019;179(5):837-841. doi:10.1002/ajmg.a.61095
89. Fiddes IT, Lodewijk GA, Mooring M, et al. Human-Specific NOTCH2NL Genes Affect Notch Signaling and Cortical Neurogenesis. *Cell*. May 31 2018;173(6):1356-1369.e22. doi:10.1016/j.cell.2018.03.051
90. Beschorner R, Waidelich J, Trautmann K, Psaras T, Schittenhelm J. Notch receptors in human choroid plexus tumors. *Histol Histopathol*. Aug 2013;28(8):1055-63. doi:10.14670/hh-28.1055
91. Dang L, Fan X, Chaudhry A, Wang M, Gaiano N, Eberhart CG. Notch3 signaling initiates choroid plexus tumor formation. *Oncogene*. Jan 19 2006;25(3):487-91. doi:10.1038/sj.onc.1209074
92. Grisanti L, Revenkova E, Gordon RE, Iomini C. Primary cilia maintain corneal epithelial homeostasis by regulation of the Notch signaling pathway. *Development*. Jun 15 2016;143(12):2160-71. doi:10.1242/dev.132704
93. Xu F, Na L, Li Y, Chen L. Roles of the PI3K/AKT/mTOR signalling pathways in neurodegenerative diseases and tumours. *Cell Biosci*. 2020;10:54. doi:10.1186/s13578-020-00416-0
94. Mirzaa G. MPPH Syndrome. In: Adam MP, Ardinger HH, Pagon RA, et al, eds. *GeneReviews(®)*. University of Washington, Seattle
Copyright © 1993-2020, University of Washington, Seattle. GeneReviews is a registered trademark of the University of Washington, Seattle. All rights reserved.; 1993.
95. Szalai R, Melegh BI, Till A, et al. Maternal mosaicism underlies the inheritance of a rare germline AKT3 variant which is responsible for megalencephaly-polymicrogyria-polydactyly-hydrocephalus syndrome in two Roma half-siblings. *Exp Mol Pathol*. Aug 2020;115:104471. doi:10.1016/j.yexmp.2020.104471
96. Dobyns WB, Mirzaa GM. Megalencephaly syndromes associated with mutations of core components of the PI3K-AKT-MTOR pathway: PIK3CA, PIK3R2, AKT3, and MTOR. *Am J Med Genet C Semin Med Genet*. Dec 2019;181(4):582-590. doi:10.1002/ajmg.c.31736
97. Mirzaa GM, Conti V, Timms AE, et al. Characterisation of mutations of the phosphoinositide-3-kinase regulatory subunit, PIK3R2, in perisylvian polymicrogyria: a next-generation sequencing study. *Lancet Neurol*. Dec 2015;14(12):1182-95. doi:10.1016/s1474-4422(15)00278-1
98. Mirzaa G, Parry DA, Fry AE, et al. De novo CCND2 mutations leading to stabilization of cyclin D2 cause megalencephaly-polymicrogyria-polydactyly-hydrocephalus syndrome. *Nat Genet*. May 2014;46(5):510-515. doi:10.1038/ng.2948
99. Fokin Artem I, Zhapparova Olga N, Burakov Anton V, Nadezhdina Elena S. Centrosome-derived microtubule radial array, PCM-1 protein, and primary cilia formation. *Protoplasma*. Sep 2019;256(5):1361-1373. doi:10.1007/s00709-019-01385-z
100. Ueki Y, Saito K, Iioka H, et al. PLOD2 Is Essential to Functional Activation of Integrin β1 for Invasion/Metastasis in Head and Neck Squamous Cell Carcinomas. *iScience*. Feb 21 2020;23(2):100850. doi:10.1016/j.isci.2020.100850
101. Neyazi B, Tanrikulu L, Wilkens L, et al. Procollagen-Lysine, 2-Oxoglutarate 5-Dioxygenase 2 Expression in Brain Arteriovenous Malformations and its Association with Brain Arteriovenous Malformation Size. *World Neurosurg*. Jun 2017;102:79-84. doi:10.1016/j.wneu.2017.02.116

102. Farajzadeh Valilou S, Alavi A, Pashaei M, et al. Whole-Exome Sequencing Identifies Three Candidate Homozygous Variants in a Consanguineous Iranian Family with Autism Spectrum Disorder and Skeletal Problems. *Mol Syndromol*. Jun 2020;11(2):62-72. doi:10.1159/000506530
103. Uribe ML, Martín-Nieto J, Quereda C, et al. Retinal Proteomics of a Mouse Model of Dystroglycanopathies Reveals Molecular Alterations in Photoreceptors. *J Proteome Res*. Jun 4 2021;20(6):3268-3277. doi:10.1021/acs.jproteome.1c00126
104. Beltrán-Valero de Bernabé D, Currier S, Steinbrecher A, et al. Mutations in the O-mannosyltransferase gene POMT1 give rise to the severe neuronal migration disorder Walker-Warburg syndrome. *Am J Hum Genet*. Nov 2002;71(5):1033-43. doi:10.1086/342975
105. Bouchet C, Gonzales M, Vuillaumier-Barrot S, et al. Molecular heterogeneity in fetal forms of type II lissencephaly. *Hum Mutat*. Oct 2007;28(10):1020-7. doi:10.1002/humu.20561
106. Hehr U, Uyanik G, Gross C, et al. Novel POMGnT1 mutations define broader phenotypic spectrum of muscle-eye-brain disease. *Neurogenetics*. Nov 2007;8(4):279-88. doi:10.1007/s10048-007-0096-y
107. Eggenschwiler JT, Anderson KV. Cilia and developmental signaling. *Annu Rev Cell Dev Biol*. 2007;23:345-73. doi:10.1146/annurev.cellbio.23.090506.123249
108. Furey CG, Choi J, Jin SC, et al. De Novo Mutation in Genes Regulating Neural Stem Cell Fate in Human Congenital Hydrocephalus. *Neuron*. Jul 25 2018;99(2):302-314.e4. doi:10.1016/j.neuron.2018.06.019
109. Beltrami B, Prada E, Tolva G, et al. Unexpected phenotype in a frameshift mutation of PTCH1. *Mol Genet Genomic Med*. Jan 2020;8(1):e987. doi:10.1002/mgg3.987
110. Delbroek H, Steyaert J, Legius E. An 8.9 year old girl with autism and Gorlin syndrome. *Eur J Paediatr Neurol*. May 2011;15(3):268-70. doi:10.1016/j.ejpn.2010.12.001
111. Ung DC, Iacono G, Méziane H, et al. Ptchd1 deficiency induces excitatory synaptic and cognitive dysfunctions in mouse. *Mol Psychiatry*. May 2018;23(5):1356-1367. doi:10.1038/mp.2017.39
112. Gavino C, Richard S. Patched1 haploinsufficiency impairs ependymal cilia function of the quaking viable mice, leading to fatal hydrocephalus. *Mol Cell Neurosci*. Jun 2011;47(2):100-7. doi:10.1016/j.mcn.2011.03.004
113. Jackson TW, Bendfeldt GA, Beam KA, Rock KD, Belcher SM. Heterozygous mutation of sonic hedgehog receptor (Ptch1) drives cerebellar overgrowth and sex-specifically alters hippocampal and cortical layer structure, activity, and social behavior in female mice. *Neurotoxicol Teratol*. Mar-Apr 2020;78:106866. doi:10.1016/j.ntt.2020.106866
114. Bisiak F, McCarthy AA. Structure and Function of Roundabout Receptors. *Subcell Biochem*. 2019;93:291-319. doi:10.1007/978-3-030-28151-9_9
115. Isogai E, Okumura K, Saito M, et al. Oncogenic Lmo3 cooperates with Hen2 to induce hydrocephalus in mice. *Exp Anim*. 2015;64(4):407-14. doi:10.1538/expanim.15-0026
116. Ramasamy SK, Lenka N. Notch exhibits ligand bias and maneuvers stage-specific steering of neural differentiation in embryonic stem cells. *Mol Cell Biol*. Apr 2010;30(8):1946-57. doi:10.1128/mcb.01419-09

117. Leblond CS, Cliquet F, Carton C, et al. Both rare and common genetic variants contribute to autism in the Faroe Islands. *NPJ Genom Med*. 2019;4:1. doi:10.1038/s41525-018-0075-2
118. Mastromoro G, De Luca A, Marchionni E, et al. External hydrocephalus as a prenatal feature of noonan syndrome. *Ann Hum Genet*. Nov 2021;85(6):249-252. doi:10.1111/ahg.12436
119. Gripp KW, Aldinger KA, Bennett JT, et al. A novel rasopathy caused by recurrent de novo missense mutations in PPP1CB closely resembles Noonan syndrome with loose anagen hair. *Am J Med Genet A*. Sep 2016;170(9):2237-47. doi:10.1002/ajmg.a.37781
120. Geoffray MM, Falissard B, Green J, et al. Autism Spectrum Disorder Symptom Profile Across the RASopathies. *Front Psychiatry*. 2020;11:585700. doi:10.3389/fpsyt.2020.585700
121. Godefroid N, Riveira-Munoz E, Saint-Martin C, Nassogne MC, Dahan K, Devuyst O. A novel splicing mutation in SLC12A3 associated with Gitelman syndrome and idiopathic intracranial hypertension. *Am J Kidney Dis*. Nov 2006;48(5):e73-9. doi:10.1053/j.ajkd.2006.08.005
122. Tsutsui H, Hamano T, Kawaura Y, et al. A case of Gitelman syndrome associated with idiopathic intracranial hypertension. *Intern Med*. 2011;50(14):1493-6. doi:10.2169/internalmedicine.50.5305
123. Nand N, Deshmukh AR, Mathur R, Chauhan V, Brijlal. Gitelman Syndrome: Presenting During Pregnancy with Adverse Foetal Outcome. *J Assoc Physicians India*. Oct 2016;64(10):104-105.
124. Narayanan R, Pirouz M, Kerimoglu C, et al. Loss of BAF (mSWI/SNF) Complexes Causes Global Transcriptional and Chromatin State Changes in Forebrain Development. *Cell Rep*. Dec 1 2015;13(9):1842-54. doi:10.1016/j.celrep.2015.10.046
125. Jin SC, Dong W, Kundishora AJ, et al. Exome sequencing implicates genetic disruption of prenatal neuro-gliogenesis in sporadic congenital hydrocephalus. *Nat Med*. Nov 2020;26(11):1754-1765. doi:10.1038/s41591-020-1090-2
126. Kerr EN, Bhan A, Héon E. Exploration of the cognitive, adaptive and behavioral functioning of patients affected with Bardet-Biedl syndrome. *Clin Genet*. Apr 2016;89(4):426-433. doi:10.1111/cge.12614
127. Cuevas E, Rybak-Wolf A, Rohde AM, Nguyen DT, Wulczyn FG. Lin41/Trim71 is essential for mouse development and specifically expressed in postnatal ependymal cells of the brain. *Front Cell Dev Biol*. 2015;3:20. doi:10.3389/fcell.2015.00020
128. Hagiwara A, Yasumura M, Hida Y, Inoue E, Ohtsuka T. The planar cell polarity protein Vangl2 bidirectionally regulates dendritic branching in cultured hippocampal neurons. *Mol Brain*. Nov 12 2014;7:79. doi:10.1186/s13041-014-0079-5
129. Lei YP, Zhang T, Li H, Wu BL, Jin L, Wang HY. VANGL2 mutations in human cranial neural-tube defects. *N Engl J Med*. Jun 10 2010;362(23):2232-5. doi:10.1056/NEJMc0910820
130. Gao B, Song H, Bishop K, et al. Wnt signaling gradients establish planar cell polarity by inducing Vangl2 phosphorylation through Ror2. *Dev Cell*. Feb 15 2011;20(2):163-76. doi:10.1016/j.devcel.2011.01.001
131. Ohata S, Alvarez-Buylla A. Planar Organization of Multiciliated Ependymal (E1) Cells in the Brain Ventricular Epithelium. *Trends Neurosci*. Aug 2016;39(8):543-551. doi:10.1016/j.tins.2016.05.004

132. Treat AC, Wheeler DS, Stolz DB, Tsang M, Friedman PA, Romero G. The PDZ Protein Na+/H+ Exchanger Regulatory Factor-1 (NHERF1) Regulates Planar Cell Polarity and Motile Cilia Organization. *PLoS One*. 2016;11(4):e0153144. doi:10.1371/journal.pone.0153144
133. Nagaoka T, Furuse M, Ohtsuka T, Tsuchida K, Kishi M. Vangl2 interaction plays a role in the proteasomal degradation of Prickle2. *Sci Rep*. Feb 27 2019;9(1):2912. doi:10.1038/s41598-019-39642-z
134. Abdel-Hamid MS, Sabry S, Abdel-Ghafar SF, El-Dessouky SH, Abdel-Salam GMH. Fetal brain arrest broadens the spectrum of WDR81-related developmental brain malformations. *Neurogenetics*. Oct 2021;22(4):287-295. doi:10.1007/s10048-021-00665-2

STROBE Statement—Checklist of items that should be included in reports of ***case-control studies***

| | Item No | Recommendation | Page No |
|---|---|---|---|
| **Title and abstract** | 1 | (*a*) Indicate the study's design with a commonly used term in the title or the abstract | 1 |
| | | (*b*) Provide in the abstract an informative and balanced summary of what was done and what was found | 1-2 |
| **Introduction** | | | |
| Background/rationale | 2 | Explain the scientific background and rationale for the investigation being reported | 4-5 |
| Objectives | 3 | State specific objectives, including any prespecified hypotheses | 5 |
| **Methods** | | | |
| Study design | 4 | Present key elements of study design early in the paper | 1-2, 6 |
| Setting | 5 | Describe the setting, locations, and relevant dates, including periods of recruitment, exposure, follow-up, and data collection | 6-7 |
| Participants | 6 | (*a*) Give the eligibility criteria, and the sources and methods of case ascertainment and control selection. Give the rationale for the choice of cases and controls | 6-7 |
| | | (*b*) For matched studies, give matching criteria and the number of controls per case | NA |
| Variables | 7 | Clearly define all outcomes, exposures, predictors, potential confounders, and effect modifiers. Give diagnostic criteria, if applicable | 7-9 |
| Data sources/ measurement | 8* | For each variable of interest, give sources of data and details of methods of assessment (measurement). Describe comparability of assessment methods if there is more than one group | 6-9 |
| Bias | 9 | Describe any efforts to address potential sources of bias | NA |
| Study size | 10 | Explain how the study size was arrived at | 6-7, Figure1 |
| Quantitative variables | 11 | Explain how quantitative variables were handled in the analyses. If applicable, describe which groupings were chosen and why | NA |
| Statistical methods | 12 | (*a*) Describe all statistical methods, including those used to control for confounding | 8-10 |
| | | (*b*) Describe any methods used to examine subgroups and interactions | NA |
| | | (*c*) Explain how missing data were addressed | 8-9 |
| | | (*d*) If applicable, explain how matching of cases and controls was addressed | NA |
| | | (*e*) Describe any sensitivity analyses | 10 |
| **Results** | | | |
| Participants | 13* | (a) Report numbers of individuals at each stage of study—eg numbers potentially eligible, examined for eligibility, confirmed eligible, included in the study, completing follow-up, and analysed | 11 |
| | | (b) Give reasons for non-participation at each stage | 11, Figure 1 |
| | | (c) Consider use of a flow diagram | 11, Figure 1 |

| | | | |
|---|---|---|---|
| Descriptive data | 14* | (a) Give characteristics of study participants (eg demographic, clinical, social) and information on exposures and potential confounders | 11 |
| | | (b) Indicate number of participants with missing data for each variable of interest | Figure 1 |
| Outcome data | 15* | Report numbers in each exposure category, or summary measures of exposure | 11, Figure 1 |

| | | | |
|---|---|---|---|
| Main results | 16 | (*a*) Give unadjusted estimates and, if applicable, confounder-adjusted estimates and their precision (eg, 95% confidence interval). Make clear which confounders were adjusted for and why they were included | 11 |
| | | (*b*) Report category boundaries when continuous variables were categorized | NA |
| | | (*c*) If relevant, consider translating estimates of relative risk into absolute risk for a meaningful time period | NA |
| Other analyses | 17 | Report other analyses done—eg analyses of subgroups and interactions, and sensitivity analyses | 12-13 |
| **Discussion** | | | |
| Key results | 18 | Summarise key results with reference to study objectives | 13-16 |
| Limitations | 19 | Discuss limitations of the study, taking into account sources of potential bias or imprecision. Discuss both direction and magnitude of any potential bias | 16 |
| Interpretation | 20 | Give a cautious overall interpretation of results considering objectives, limitations, multiplicity of analyses, results from similar studies, and other relevant evidence | 17-18 |
| Generalisability | 21 | Discuss the generalisability (external validity) of the study results | 17 |
| **Other information** | | | |
| Funding | 22 | Give the source of funding and the role of the funders for the present study and, if applicable, for the original study on which the present article is based | 18 |

*Give information separately for cases and controls.

**Note:** An Explanation and Elaboration article discusses each checklist item and gives methodological background and published examples of transparent reporting. The STROBE checklist is best used in conjunction with this article (freely available on the Web sites of PLoS Medicine at http://www.plosmedicine.org/, Annals of Internal Medicine at http://www.annals.org/, and Epidemiology at http://www.epidem.com/). Information on the STROBE Initiative is available at http://www.strobe-statement.org.
